# Transcriptomic and genetic analysis suggests a role for mitochondrial dysregulation in schizophrenia

**DOI:** 10.1101/2025.03.14.25323827

**Authors:** Lisa Bast, Shuyang Yao, José A. Martínez-López, Fatima Memic, Hayley French, Milda Valiukonyte, Robert Karlsson, Jia Wen, Jie Song, Ruyue Zhang, Anthony Abrantes, Frank Koopmans, Anne-May Österholm, Gorazd Rosoklija, J. John Mann, Aleksandar Stankov, Iskra Trencevska, Andrew Dwork, Craig A. Stockmeier, Michael I. Love, Paola Giusti-Rodriguez, August B. Smit, Patrick F. Sullivan, Jens Hjerling-Leffler

## Abstract

Schizophrenia is an often devastating disorder characterized by persistent and idiopathic cognitive deficits, delusions and hallucinations. Schizophrenia has been associated with impaired nervous system development and an excitation/inhibition imbalance in the prefrontal cortex. On a molecular level, schizophrenia is moderately heritable and genetically complex. Hundreds of risk genes have been identified, spanning a heterogeneous landscape dominated by loci that confer relatively small risk. Bioinformatic analyses of genetic associations point to a limited set of neurons, mainly excitatory cortical neurons, but other analyses suggest the importance of astrocytes and microglia. To understand different cell type roles in schizophrenia and reveal novel cell-type specific aetiologically relevant perturbations in schizophrenia, our study integrated genetic analysis with single nucleus RNA-seq of 536,618 nuclei from postmortem samples of dorsal prefrontal cortex (Brodmann Area 8/9) of 43 cases with schizophrenia and 42 neurotypical controls. We found no significant difference in cell type abundance. Gene expression in excitatory layer 2-3 intra-telencephalic neurons had the greatest number of differentially expressed transcripts and, together with excitatory deep layer intra-telencephalic neurons, conferred most of the genetic risk for schizophrenia. Most differential expression of genes was found in specific cell types and was dominated by down-regulated transcripts. Down-regulated transcripts were enriched in gene sets including transmembrane transport, mitochondrial function, protein folding, and cell-cell signaling whereas up-regulated transcripts were enriched in gene sets related to RNA processing, including RNA splicing in neurons. Co-regulation network analysis identified 40 schizophrenia-relevant programs across 13 cell types. A gene program largely shared between neuronal subtypes, astrocytes, and oligodendrocytes was significantly enriched for schizophrenia risk, supporting an aetiological role for perturbed protein modification, ion transport, and mitochondrial function. These results were largely consistent with cell-type expression quantitative trait locus and transcriptome-wide association analyses. Moreover, single-cell RNA sequencing results, most prominently mitochondrial dysfunction, had multiple points of convergence with proteomic and long-read RNA sequencing results from samples from the same donors. Our study integrates genetic analysis with transcriptomics to reveal novel cell-type specific aetiologically relevant perturbations in schizophrenia.

## Introduction

Schizophrenia is a common and highly heritable psychiatric disorder often accompanied by substantial impairment of quality of life, mortality, and personal or societal costs (Saha et al. 2005; Saha, Chant, and McGrath 2007; “Global, Regional, and National Burden of 12 Mental Disorders in 204 Countries and Territories, 1990-2019: A Systematic Analysis for the Global Burden of Disease Study 2019” 2022; Knapp, Mangalore, and Simon 2004).

We recently reviewed the epidemiology, genetic epidemiology, and genomics of schizophrenia (Patrick F. Sullivan, Yao, and Hjerling-Leffler 2024), Supplemental Information, Section 1), and several points stood out. First, the progression of knowledge has been orderly, with the initial findings for rare copy number variation, exome sequencing, and genome-wide association studies continuing to be supported by subsequent studies. Second, because of the size and rigor of the published studies (Purcell et al. 2009; Singh et al. 2016; Trubetskoy et al. 2022; Singh et al. 2022; Marshall et al. 2017), the number of variants, their frequencies in schizophrenia cases and neurotypical controls, and the degree of risk conferred are clearer now than ever before. Third, schizophrenia is highly polygenic: to date there are >300 single nucleotide polymorphisms (SNPs) in the form of common variation (287 genomic regions), rare variation in protein-coding regions of 10 genes, and 13 rare copy number variants. Even the strongest findings are rare in schizophrenia cases: e.g., *SETD1A* has impactful exonic variants in 0.12% of cases and 0.017% of controls, and the 22q11.2 deletion is found in 0.30% of cases and 0.0049% of controls.

Following the central dogma of biology, it is natural to combine genomics with transcriptomics. Previously, most published studies analyzing RNA from postmortem sections of cortex in bulk, involved laser microdissection (Harris et al. 2008) or isolation of cells based on markers (e.g. Gandal, Haney, et al. 2018). Recent advances enable single-nucleus RNA-seq to study the cellular complexity of the human brain at much higher resolution. The Human Brain Atlas paper (Siletti et al. 2023) reported between 461 and 3313 identifiable cell types arranged in 31 superclusters. Due to the cellular complexity of the brain, understanding transcriptomic changes in schizophrenia requires cellular resolution. In prior studies (Skene et al. 2018; Bryois et al. 2020; Yao et al. 2024), we estimated the SNP-heritability enrichment of schizophrenia in brain cell types. In all papers, enrichment was only found in neuronal cell-types, and non-neuronal cells did not show enrichment. Our most recent papers (Skene et al. 2018; Bryois et al. 2020; Yao et al. 2024) used the Human Brain Atlas data with more complete brain coverage (Siletti et al. 2023). We found significant etiological relations of schizophrenia to 14 neuronal superclusters which pointed to the cortex, amygdala, and hippocampus as key brain regions to investigate.

Here we extend our previous enrichment findings and report results of snRNA-seq on the prefrontal cortex from 43 cases with schizophrenia and 42 controls. We test for cell type abundance changes and identify genes and gene networks that differ between cases and controls in the cell types of the prefrontal cortex. Moreover, we identified schizophrenia genetic risk enriched co-expression modules across cell types and functions and assessed the overlap of findings across different levels of analyses: differential gene expression, co-expression network analysis, transcriptome wide association study (TWAS), and proteomics (Koopmans et al.) and long-read RNA-seq (Abrantes et al.) results from the same samples. We combine these results with genome-wide genotypes to confirm an aetiological role for excitatory neurons and changes in mitochondria and identify RNA-processing and synaptic dysfunction as central processes downstream of the disorder.

## Results

### Single nucleus RNA-sequencing and genotyping of 85 prefrontal cortex donor samples

We analyzed postmortem frozen human tissue from BA8/BA9 of 43 schizophrenia samples and 42 age-, postmortem interval (PMI)-, and sex-matched controls using snRNA-seq (Fig. 1A; Table T1). We genotyped all samples with a SNP array (Illumina GSA). There was one large CNV identified in a control (96% overlap with 16p11.2 deletion CNV) and no large CNV identified in cases. We did not identify any significant difference in CNV burden between cases and controls (Table T11). Of 83 samples (41 cases and 42 controls) that passed quality control for snRNA-seq (Fig. S1A-F), 73 samples (36 cases and 37 controls) were of European ancestry (Fig. 1B). As expected, we observed significantly higher (p = 0.0034) polygenic schizophrenia risk scores in cases compared to controls (Fig. 1C, Table T1, (Trubetskoy et al. 2022).

**Figure 1.**
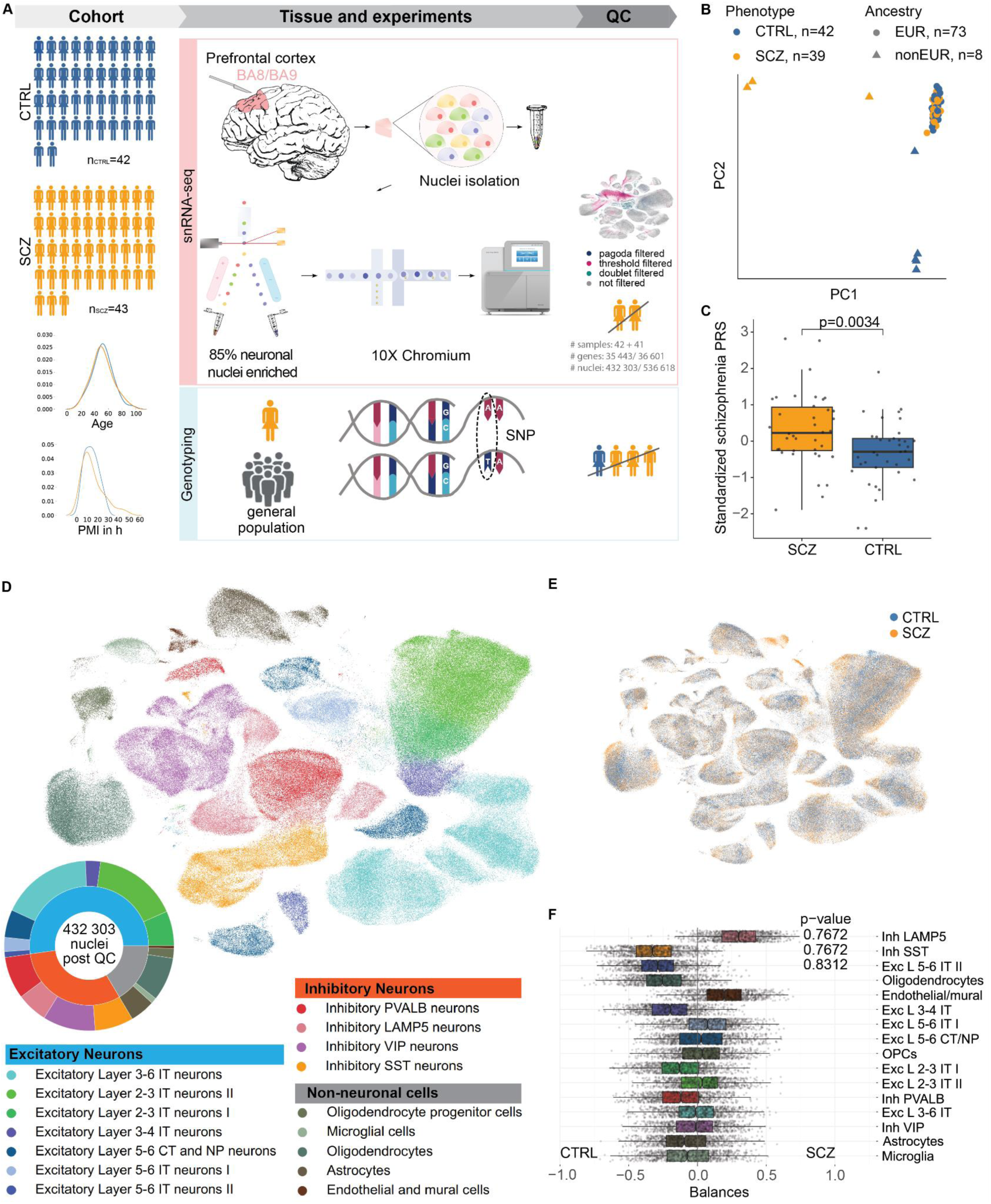
**(A)** Study summary. Both snRNA-sequencing and genotyping were performed on tissue from the prefrontal cortex (Brodmann area 8 or 9) of 43 schizophrenia (SCZ) and 42 control group (CTRL) samples. Differential abundance analysis was performed on cell type annotated snRNA-seq data and polygenic risk scores calculated from individual genotypes. (**B)** First and second principal component (PC) of genotyping data containing 81 samples that passed quality control in both snRNA-sequencing and genotyping. The cluster to the top-right corner corresponds to donors with European ancestry (n=73). (**C)** Standardized polygenic risk scores (PRS) restricted to European ancestry were significantly (t-test, p-value = 0.0034) higher in SCZ than in CTRL. Center lines of box plots correspond to median, box limits to upper and lower quartiles, and whiskers to 1.5x interquartile range. The points correspond to individual PRS values. (**D)** Uniform manifold approximation and projection (UMAP) of single nuclei with the cell type identity for 16 clusters. Donut chart indicating number of nuclei per cell type (16 clusters, outer ring) and broader cell type class (3 clusters, inner ring). (**E)** UMAP of single nuclei post quality control (QC) highlighting disease status. (**F)** Differential abundance testing of 16 cell type clusters reveals non-significant differences in cell type abundance in schizophrenia patients. Cell types are sorted by their p-values resulting from two-sided testing if their balances significantly deviate from zero. The smallest adjusted p-values (0.7672, inhibitory LAMP5 and SST cells) were non-significant. Center lines of box plots correspond to median, box limits to upper and lower quartiles, and whiskers to 1.5x interquartile range. The points correspond to individual values for resampled balances.

Preprocessed snRNA-seq data (Fig. 1D-E) were integrated and semi-automatically annotated (Figs. S2-S3, Supplemental Methods) to group them into 3 broad classes and 16 cell types (Fig. 1D, Tables T2-T3). Differential cell type abundance testing using cacoa (Petukhov et al. 2022) revealed no significant changes between cases and controls for any of the 16 cell types (Fig. 1F).

### Excitatory intra-telencephalic neurons have most DEGs and are enriched for common genetic variants associated with schizophrenia

Gene expression analysis using a DESeq2 pseudo-bulk approach (Love et al. 2014, Fig. 2A) revealed that differentially expressed genes (DEGs, q-value < 0.05) were predominantly found in the excitatory clusters, most notably in superficial layer excitatory intra-telencephalic (IT) neurons (Fig. 2B,C). As expected, there was greater power to detect DEGs with higher cell number (Fig. 2D). Although non-neuronal cells and inhibitory neurons tended to have fewer DEGs, some smaller excitatory cell type populations had more DEGs than expected purely from a linear power increase. Thus, the enrichment of DEGs in excitatory cells was not solely driven by the number of cells in the clusters. DEGs were predominantly down-regulated (Fig. 2C), as noted in a prior snRNA-seq study (Ruzicka et al. 2024). Spearman correlation analysis of DEG’s signed adjusted p-values between cell types revealed a remarkable specificity of DEGs between cell types (Spearman’s rho < 0.3, Fig. 2E, Fig. S6D-E). Schizophrenia SNP-heritability enrichment analysis (Finucane et al. 2015) of the top 10% most specifically expressed (top decile expression proportion: TDEP) genes in each cell type revealed that the common genetic basis of schizophrenia was enriched in six excitatory cell types (Fig. 2F). This includes superficial layer excitatory IT types that also had the most altered expression (Fig. 2C).

**Figure 2.**
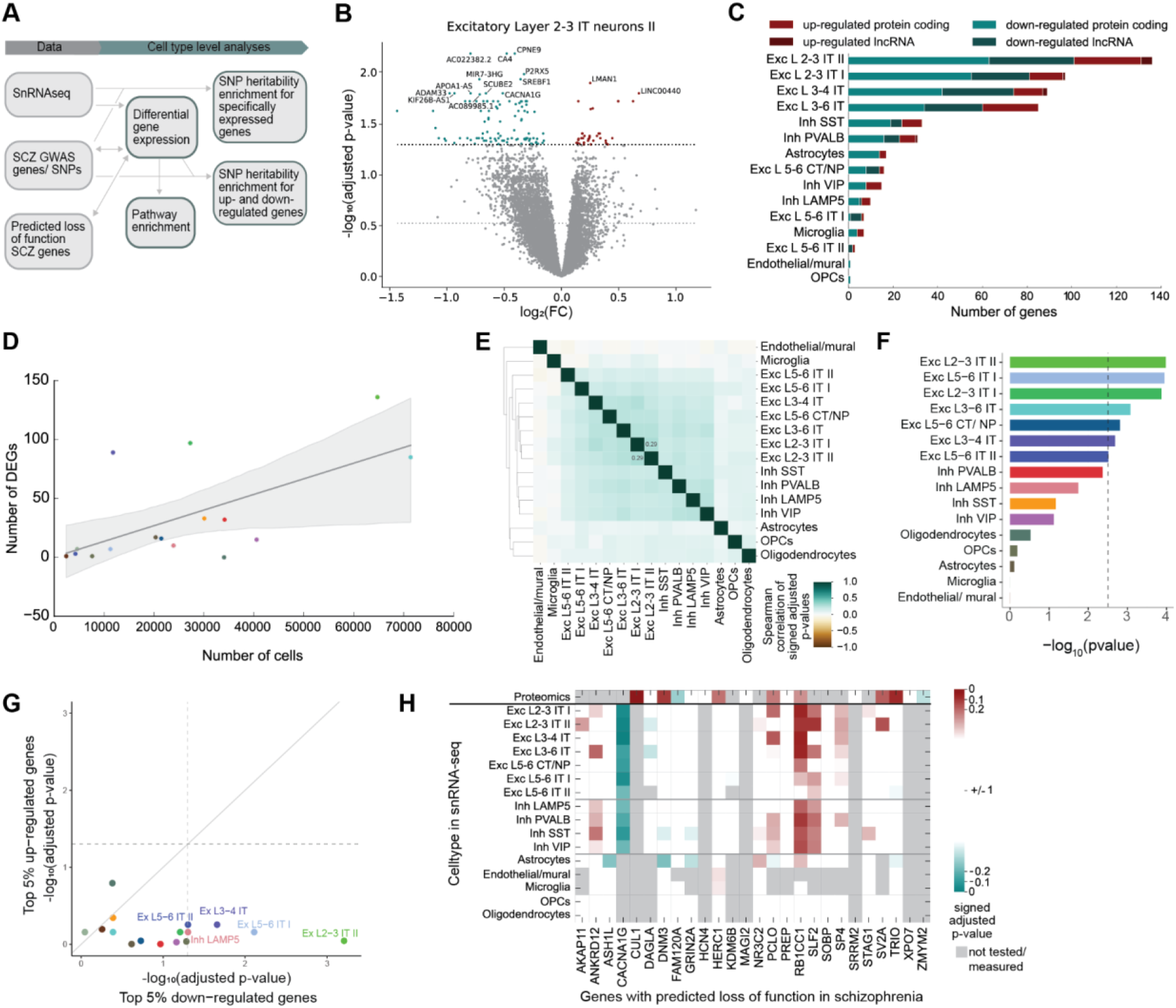
**(A)** Data overview and analysis summary. Double-headed arrows indicate comparison of results or datasets. (**B)** Volcano plot for excitatory layer 2-3 IT neurons II highlighting genes up- (red) and down-regulated (green) in schizophrenia (dashed lines, alpha=0.05 and alpha=0.3). **(C)** Number of up- and down-regulated genes in schizophrenia identified per cell type cluster (left) are highest in several excitatory neurons and lowest in oligodendrocyte and endothelial lineage. Most up-regulated genes are protein-coding. **(D)** Top five cell types with the highest number of DEGs have a low, medium and high number of cells. Cell types containing many cells obtain not necessarily a high number of DEGs. **(E)** Cell type specificity of DEGs highlighted by low (rho <0.3) Spearman correlation values of signed p-values in DEG analysis between each pair of cell types. Only a few DEGs are shared between inhibitory and excitatory cell types, respectively. **(F)** Estimated enrichment of SCZ SNP-heritability in the top 10% specifically expressed genes per cell type (right). Six excitatory cell types are significantly enriched for SCZ SNP-heritability (dashed line, alpha=0.05, Bonferroni-corrected). **(G)** P-values in enrichment analysis in the 5% most down- and up-regulated genes for each cell type. The most down-regulated genes are more significantly enriched for SCZ SNP-heritability than the most up-regulated genes. Down-regulated genes from the seven labeled cell types are significantly enriched for SCZ SNP-heritability with FDR≤0.05. **(H)** Genes with predicted loss of function in SCZ (Singh et al. 2022) and their signed adjusted p-value in differentially abundant protein (DAP) analysis (first row), and DEG analysis across cell types (remaining rows).

To evaluate the relationship of these results to schizophrenia risk in an orthogonal manner, we identified DEGs in excitatory neurons, inhibitory neurons, and non-neuronal cells and evaluated their relation to genes in GWAS loci for schizophrenia, bipolar disorder, major depression, IQ, educational attainment and neuroticism (Trubetskoy et al. 2022; Mullins et al. 2021; Howard et al. 2019; Okbay et al. 2022; Savage et al. 2018). Notably, there were only a few DEGs in the extended Major Histocompatibility Complex (MHC) region. Excitatory DEGs were more likely to be in a schizophrenia GWAS locus (16.9% vs 9.4%, P = 7e-4). There were no significant differences for any other GWAS trait or for any other cell types. These findings suggest a relatively specific relationship of schizophrenia GWAS findings and DEGs in excitatory neuronal nuclei from cases compared with controls.

### Common genetic variants are generally down-regulated while predicted loss of function genes are mostly up-regulated in neurons

We then separately investigated up- and down-regulated genes regarding their relationship with schizophrenia common and rare genetic variants and performed SNP-heritability enrichment analysis of the top 5% up- and down-regulated genes for each cell type. Overall genes with decreased expression had greater enrichment (Fig. 2G). Specifically, the 5% most down-regulated genes in five excitatory IT types, inhibitory somatostatin (SST) neurons, and oligodendrocytes (highlighted in figure Fig. 2G) were significantly enriched for schizophrenia SNP heritability. Our interpretation is that common schizophrenia-risk variants are more likely to reduce gene expression in a disease state (top 5% Fig. 2G, other 5% bins Fig. S7). The predicted loss of function mutation schizophrenia genes (Singh et al. 2022) with large effect size however, with the important exception of CACNA1G, tended to be more up-regulated (Fig. 2H), suggesting that these classes of genes might have different roles in schizophrenia pathophysiology.

### Pathway analysis of DEGs highlights up-regulated RNA processing and diverse down-regulated processes including mitochondrial function

We next performed GO-term gene set enrichment analysis to identify biological pathways in the DEG results. As the sample size and consequently the number of DEGs per cell type is limited, we included genes with adjusted p-values < 0.3 in the DEG analysis (14,151 DEGs across 16 cell types, Fig. S6, Table T5) when performing pathway enrichment analysis.

Overall, among down-regulated genes we observed transmembrane transport, GTPase related pathways, energy-related gene ontology (GO) terms (mitochondrion organization, mitochondrial electron transport, ATP-related processes), protein folding, cell-cell signaling, and cell migration. For up-regulated DEGs, pathways were predominantly RNA processing (including splicing), and chromatin organization (Fig. 3A, S8-9). Cytoplasmic translation and neuron differentiation pathways were enriched for both, up- and down-regulated DEGs (Fig. 3A). Additional gene set analyses revealed enrichment for genes with binding sites for RNA-binding proteins (RBFOX1-3, CELF4, and IMP2), higher conservation (gnomAD pLI), and overlap with cortical bulk-tissue schizophrenia case/control DEGs (Gandal, Zhang, et al. 2018, Table T6).

**Figure 3.**
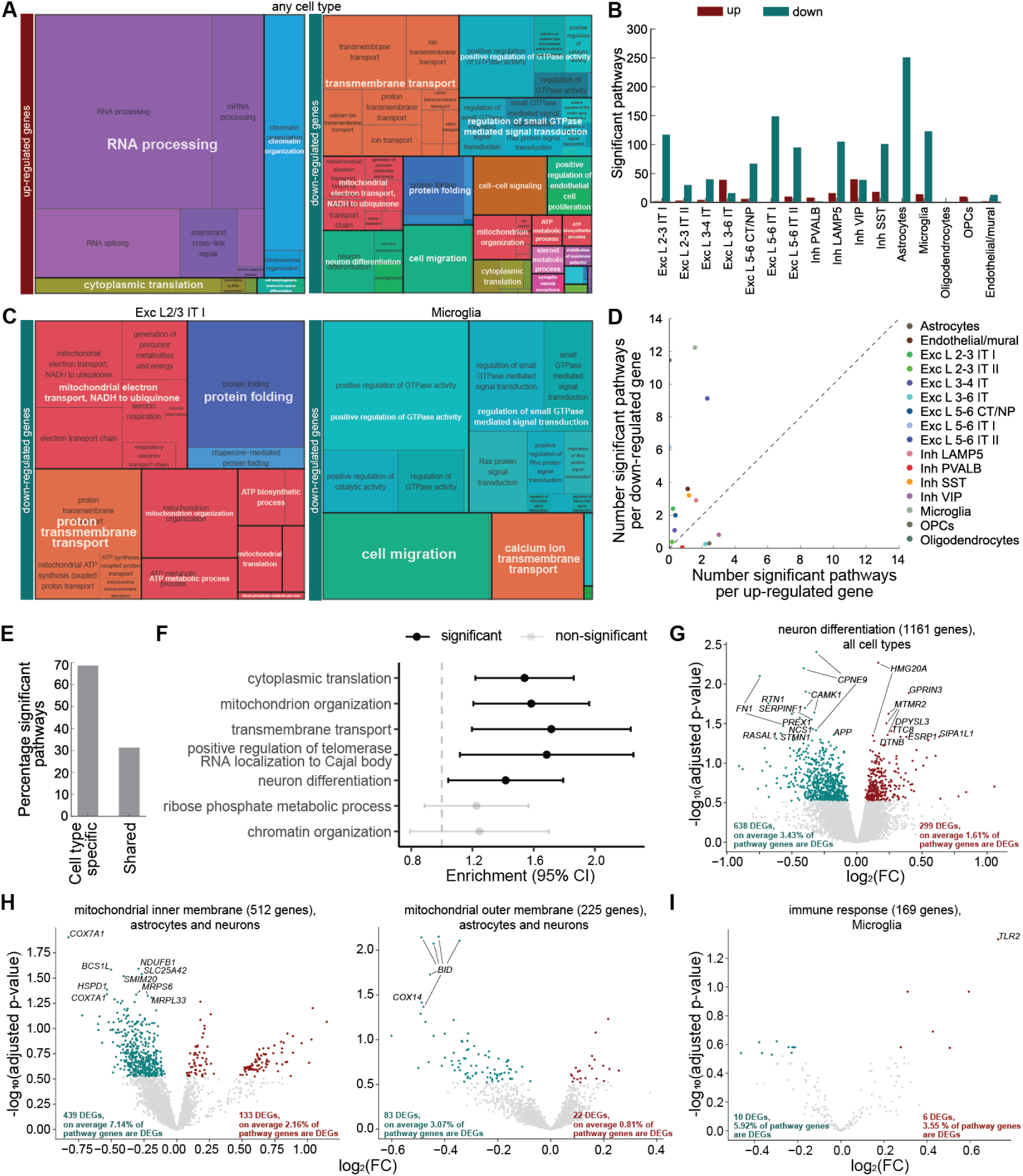
**(A)** Significantly up- (left) and down-regulated (right) pathways of GO biological processes (BP) pathways in at least one cell type restructured into broad functional categories with rrvgo. RNA processing and chromatin organization are identified to be up-regulated and mitochondrial function, neuron development, transmembrane transport, cell-cell signaling, GTPase activity and protein folding to be down-regulated. Cytoplasmic translation is both up-and down-regulated. **(B)** Number of significant GO term pathways for each cell type’s up- and down-regulated genes (DEG analysis alpha=0.3). **(C)** Significantly down-regulated pathways of GO BP pathways specifically for two cell types, showing impaired mitochondrial pathways, protein folding and proton transmembrane transport in excitatory layer 2-3 IT I neurons (left) and cell migration, GTPase mediated signal transduction and calcium ion transmembrane transport in microglia (right). **(D)** Average number of significant pathways per up- and down-regulated gene for each cell type identifying whether pathways are perturbed through down-regulation (i.e. astrocytes) or up-regulation (i.e. inhibitory PVALB neurons) or both (inhibitory LAMP5 neurons). **(E)** Percentage of significant pathways are primarily cell type specific. **(F)** DEG (alpha≤0.3) in five pathway categories show significant enrichment for common SCZ risk. Error bars indicate the 95% confidence interval. **(G)** Volcano plot of neuron differentiation genes in any cell type shows twice as many down-regulated than up-regulated genes. **(H)** Volcano plot of mitochondrial inner (left) and outer (right) membrane GO term genes comprising differential gene expression analysis results of astrocytes and neuronal cell types show down-regulation of inner mitochondrial membrane is strongest. **(I)** Volcano plot of immune response genes in microglia shows stronger down-than up-regulation.

### Shared and distinct pathways across cell types

For each cell type, the GO-term analysis of DEGs revealed a detailed level of organization (Fig. 3B, Fig. S8-9, Table T6). In most cell types, the number of significant gene sets per down-regulated DEG was higher than the number per up-regulated DEG (Fig. 3D). Most significant gene sets were cell type-specific, and only 30% were shared between cell types (Fig. 3E; Fig. S12A). Mitochondrial function emerged as a common GO term amongst down-regulated pathways common to 9 of 11 neuronal cell types (with the exception of excitatory L3-6 and L5-6 IT type II neurons) and astrocytes. Protein-folding gene sets showed a similar pattern. For up-regulated DEGs, we found gene set enrichments for mRNA metabolism and splicing in 6 of 11 neuronal cell types. These terms were not enriched in non-neuronal cells except for oligodendrocyte progenitor cells (OPCs). As a confirmation we compared these DEGs and pathways to differential isoform usage genes and their pathways, measured with long-read RNA-sequencing in a subset of our samples (Table 1, Abrantes et al.). In addition to several synaptic pathways, this analysis revealed that the spliceosomal complex itself was significantly enriched for differential isoform usage genes (Fig. S13C). Differential isoform usage genes and differentially expressed genes were however mostly non-overlapping (Fig. S13A-B).

We also identified pathways altered in specific cell types (Fig. S8-10, Table T6). Excitatory L2-3 IT type I neurons had an additional set of mitochondrial enrichments including cytochrome, proton- and electron-transport as well as ATP-related terms. Similarly, L5-6 IT type I and II neurons showed complementary enrichments where type I had more enrichments for synapse-related terms (along with astrocytes), astrocytic development and lipid transport, and type II instead were enriched for terms related to cell growth and adhesion to extracellular matrix/substrate. For inhibitory neurons we also observed specific signals where LAMP5 cells had the most down-regulated enrichments including synaptic signaling, cell growth and terms related to epithelial and endothelial cell migration/proliferation, and SST and VIP neurons had the most up-regulated pathways (DNA repair and DNA replication). PVALB cells had no significant enrichments beyond RNA processing and mitochondrial pathways that were shared by most neurons. SynGO analysis (Koopmans et al. 2019) suggested that the impaired synaptic function of excitatory deep layer intra-telencephalic and inhibitory SST neurons coalesced on genes contributing to both pre- and postsynaptic pathways (Fig. S11).

Exclusively in microglial cells, we find several GTPase mediated signal transduction pathways of the ras and rho superfamilies to be down-regulated (Fig. 3C, S8C, S9A), which regulate cellular events such as proliferation, differentiation, or apoptosis, cellular growth, metabolism and gene expression (Sastre et al. 2020). In accordance with this we saw an up-regulation in mitochondria and translation genes suggesting increased biogenesis. We also observed a down-regulation of cell migration in microglia. OPCs had neuronal development GO terms and splicing up-regulated and cell migration terms down.

### SNP enrichment points to possible causal role for changes in cytoplasmic translation, mitochondrion organization, transmembrane transport and neuron differentiation

To investigate a possible causal role for the enriched pathways, we sought to identify pathways enriched for common schizophrenia risk and performed a schizophrenia SNP-heritability enrichment analysis of the genes included in each GO-term. Out of 22 GO biological process pathway categories (Fig. S9A), 7 contained sufficient (>1%) linkage disequilibrium score regression (LDSC) SNPs to perform the analysis, and 5 showed significant enrichment of common genetic risk of schizophrenia (Fig. 3F, S12B), including cytoplasmic translation, mitochondrion organization, transmembrane transport, positive regulation of telomerase RNA localization to Cajal body, and neuron differentiation. The latter three were also confirmed by pathway enrichment of mBATcombo-significant genes, i.e. genes significantly enriched for gene-level genetic risk associated with schizophrenia (Fig. S12D, A. Li et al. 2023, Supplemental Methods). Many neuron differentiation (GO:0030182) DEGs were down-regulated across several cell types (CPNE9, CAMK1, FN1, and SERPINF1), while MTMR2 and HMG20A were up-regulated in three and two cell types, respectively (Fig. 3G). Interestingly, we found more DEGs from mitochondrial inner membrane (GO:0005743) than outer membrane (GO:0005741, Fig. 3H), suggesting that the mitochondrial dysfunction is in large part attributable to dysfunction in the inner mitochondrial membrane. In microglial cells, the immune response (GO:0006955) pathway was significantly enriched for down-regulated DEGs (adjusted p-value = 0.03, Table T6). In addition, we identified TLR2, important for pathogen recognition and activation of innate immunity, as a highly significant and large effect size up-regulated immune response pathway gene (Fig. 3I).

### TWAS based on cell type-level eQTLs supports central transcriptomic findings

To prioritize schizophrenia risk genes based on transcriptome and genotype data, we first performed cell type-level expression quantitative trait loci (eQTL) analysis, and then transcriptome-wide association analysis (TWAS, (Yang, Yeung, and Liu 2021). For eQTL analysis, we identified 1,122, 909, and 327 genes in the significant eQTL pairs (referred as eGenes in the following text) for excitatory neuronal, inhibitory neuronal, and non-neuronal classes at a 5% false discovery rate (FDR). A total of 4,357 eGenes were identified in at least one cell type (Fig. 4A, Supplementary Table T10). As expected, the number of identified eGenes was positively correlated with the number of cells in the cell type (Pearson’s rho = 0.82, p = 9.86e-05, Fig. 4B), and SNPs in the significant eQTL pairs (referred as eSNPs) are distributed near the transcription start site (Fig. 4C). The effect size of the eQTL pairs correlated with the fully mapped bulk-based eQTL pairs (GTEx brain region BA9, Pearson’s rho = 0.89, Fig. 4D) and previously reported eQTLs at a cell class level (Bryois et al. 2022), Pearson’s rho = 0.87, Fig. 4E). The eQTL effect size was also highly correlated between cell classes (Pearson’s rho > 0.8, Fig. 4F). Overlaying the eSNPs with human brain cell type-specific candidate cis-reculatory elements (cCRE, (Y. E. Li et al. 2023) showed enrichment of the eSNPs in the cell type cCREs (Fig. 4G), reflecting the potential genetic regulatory role of some eSNPs. Interestingly, eSNPs at the cell type level had higher specificity of the enrichment in the cCREs than the eSNPs at the cell class level; for instance, the enrichment of excitatory layer 2-3 IT neurons cCREs was more profound around eQTLs of specific excitatory cell types (Fig. 4H) than the class-level eQTLs of the excitatory neurons (Fig. 4G). Similarly, oligodendrocytes cCREs were specifically enriched around the oligodendrocytes eQTLs (Fig. 4I), while the enrichment was not obvious for the non-neuronal cell class. These observations support a cell type-specific regulatory function of eSNPs.

**Figure 4.**
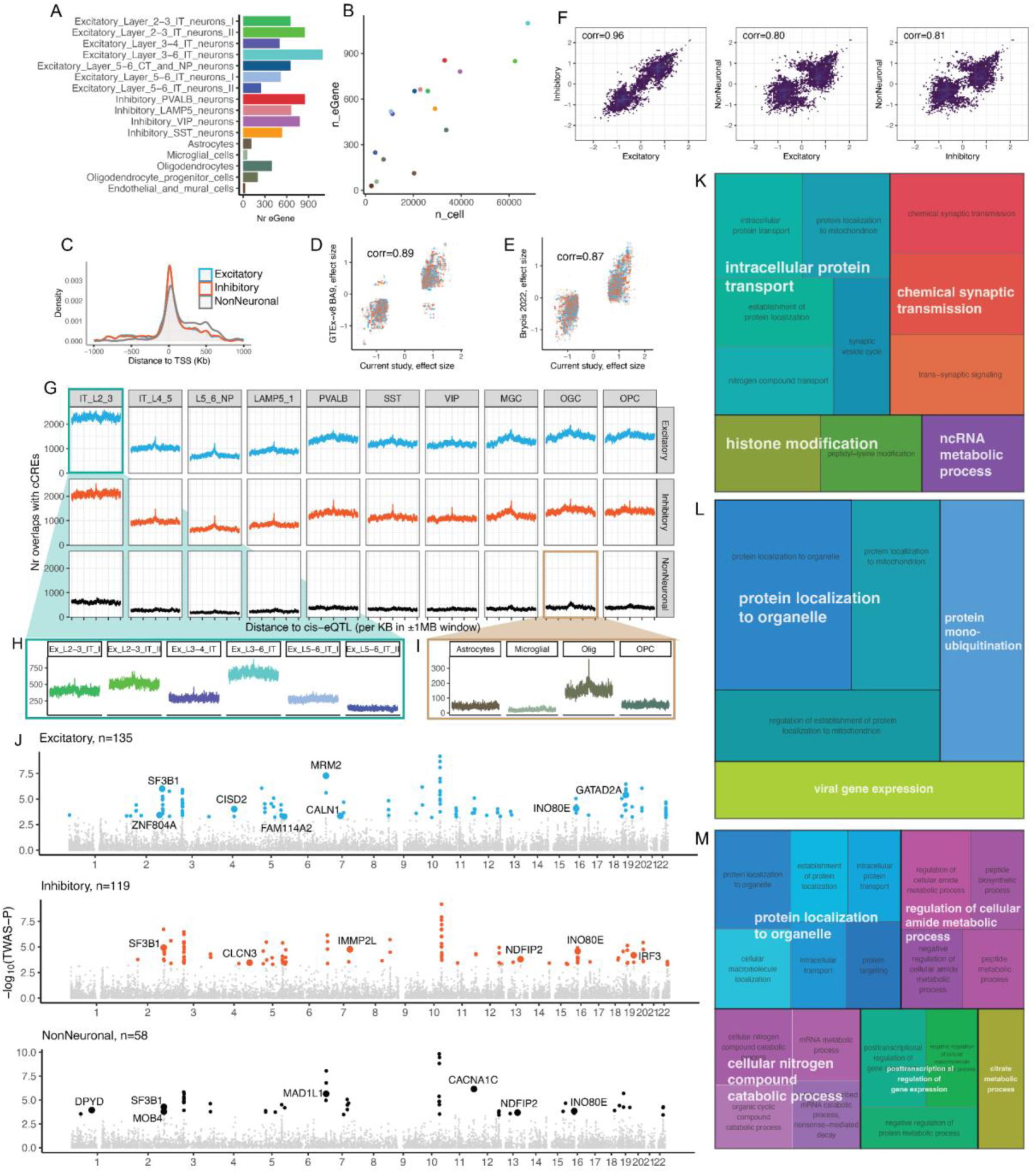
**(A)** Number of eGenes (genes with any eQTL at FDR≤0.05) per cell type. **(B)** Number of eGenes are correlated with the number of cells per cell type. **(C)** eQTLs are enriched around the transcription start site (TSS). **(D-E)** The effect size of the cell class eQTL showed high correlation with previous eQTLs based on bulk RNA-sequencing from GTEx (v8) brain region BA9 **(D)** and cell type eQTL findings from Bryois et al., 2022 **(E)**. **(F)** eQTLs have high correlation in effect size between different cell classes, especially the neuronal cell classes. **(G)** cCREs are enriched near the cell class eQTLs. The horizontal names are the cCRE cell types, and the vertical names are the eQTL cell class names. **(H)** The cCREs in layer 2-3 IT neurons were enriched around eQTLs of specific excitatory cell types. The name (horizontal) of each plot corresponds to the eQTL cell type. **(I)** Oligodendrocytes cCREs were specifically enriched around the oligodendrocyte eQTLs. The name (horizontal) of each plot corresponds to the eQTL cell type. **(J)** TWAS significance in three cell classes: excitatory neurons, inhibitory neurons, and non-neuronal cells. Each dot in the plot represents a gene; colored dots indicate genes with TWAS-FDR≤0.05, and enlarged dots with labels of gene names are the TWAS-FDR≤0.05 genes that were also prioritized in the latest GWAS (Trubetskoy et al. 2022). The full list of TWAS-FDR≤0.05 genes are listed in Supplementary Table T13. TWAS significance at cell type level is presented in Supplementary Figure S15-16. **(K-M)** Treeplot of GO:BP pathways enriched in TWAS-FDR≤0.05 genes for cell types belong to the excitatory neurons **(K)**, inhibitory neurons **(L)**, and non-neuronal **(M)** classes. The GO pathways are detailed in Supplementary Figure S18.

We then performed TWAS integrating schizophrenia GWAS (Trubetskoy et al. 2022) with our cell class and cell type eQTLs to nominate genes. Our TWAS analysis implicated 228 genes at a cell class level and prioritized 16 genes from the GWAS (Fig. 4J), while the cell type level analysis identified 582 genes including 35 genes in the GWAS (Fig. S14-S17). GO terms enriched in the cell type level TWAS genes showed agreement with those enriched in the DEGs, including RNA processing, intracellular protein transport and localization, and regulation of gene expression (Fig. 4K-M). Interestingly, mitochondria related pathways are specific to neuronal cell types, including excitatory layer 3−4 IT neurons, and LAMP5, SST, and VIP interneurons while mRNA-binding is observed in a broader range of cell types (Fig. S18).

### Energy supply pathways co-occur with protein-modification, protein-assembly and impaired ion-transport in schizophrenia-relevant gene program in several cell types

To investigate which gene co-expression programs were perturbed, we applied high dimensional weighted gene co-expression network analysis (Morabito et al. 2023). For each cell type we analyzed the 2,000 most variable genes revealing gene co-expression modules (Fig. 5A). We excluded microglial, endothelial and mural cells as well as excitatory layer 5-6 IT II neurons due to insufficient numbers of cells and/or donors contributing to these cell types (Supplemental Methods). 40 of the 92 identified modules were significantly altered in cases versus controls (Fig. 5B). Notably, the most significant module (”red”, excitatory layer 2-3 IT neurons II) contained four hub genes coding for proteins identified as significantly higher (TUBA1B) and lower (HOMER1, NPTX2, and VGF) abundant in schizophrenia in the proteomic analyses of overlapping samples (Fig. 5C, Table T1, Koopmans et al.). This module was related to neuron differentiation and apoptosis, response to peptide, regulation of transport and localization, but also pre- and post-synaptic pathways, synaptic signaling and neuronal dense core vesicle SynGO terms (Fig. 5D). We performed pathway enrichment analysis of all schizophrenia-relevant modules’ genes (Fig. 5E, S19D, Table T7) and grouped similar pathways into categories. Certain modules contained specific pathways and mapped only to a few categories (e.g. excitatory layer 5-6 CT/NP pink module - synapse function and OPCs black module - cell-cell signaling/nervous system development). Other modules contained pathways across many different categories, such as astrocytes turquoise and excitatory layer 2-3 II yellow. 10 modules were not significantly enriched for any pathway (Fig. 5E), partly due to their rather small module sizes (Fig. 5F). Next, we analyzed the overlap of these modules with respect to the genes they contain and identified 2 groups with substantial gene overlap in 8 and 5 different cell types, which we named the “bordeaux” and the “midnight” modules respectively (Fig. 5F). Interestingly, all 8 bordeaux modules’ genes were significantly enriched for protein-modification, Energy (ATP) pathways, extracellular matrix, and ion transport (Fig. 5E), suggesting a largely similar program is perturbed in inhibitory VIP, PVALB and LAMP5 neurons, superficial and deep layer neurons, astrocytes and oligodendrocytes. Despite relatively high gene-overlap (Fig. 5F), the midnight modules’ pathways were rather specific with only minor overlap in terms of GO-enrichment (Fig. 5E). Across the modules significantly changed in schizophrenia, most genes were classified as nuclear mitochondrial genes, RNA binding protein coding genes, or other protein coding genes (Fig. S20D).

**Figure 5.**
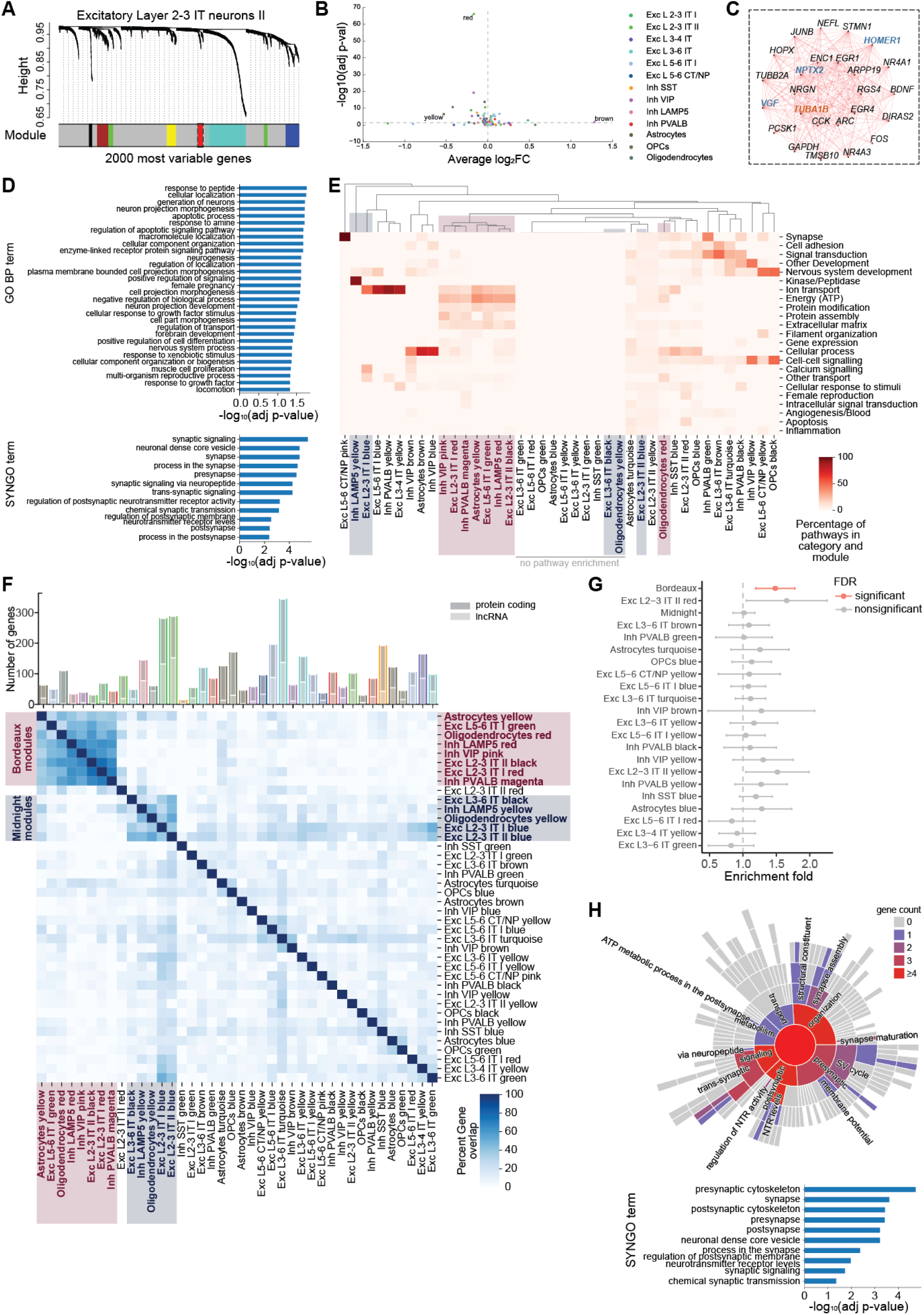
**(A)** Dendrogram tree for excitatory layer 2-3 IT neurons II with module affiliation for each of the 2000 most variable genes. **(B)** 40 modules were identified as differentially expressed in schizophrenia (DEMs) across 13 cell types. **(C)** Hub gene network for excitatory layer 2-3 IT neurons II red module with genes highlighted in blue/orange are coding for proteins with significantly lower/higher abundance in SCZ. **(D)** Top 30 GO BP terms (top) and significantly enriched SYNGO terms (bottom) with their −log_10_(adjusted p-values) for the excitatory layer 2-3 IT neurons II red module genes. **(E)** Functional specificity of DEMs as percentage of GO BP pathways per category and module showing shared functions across cell type-specific modules. 10 modules were not enriched for any pathway. The 8 bordeaux modules are mostly Ion transport, Energy (ATP), Protein modification and extracellular matrix pathways. The 5 midnight modules are functionally rather distinct despite their gene overlap and only obtain nervous system development as a common category. **(F)** Number of genes in each cell type specific DEM (top) and similarity of modules as percentage of genes overlapping with other modules (bottom) identified a group of 8 and 5 modules with substantial gene overlap (bordeaux and midnight modules). **(G)** Mean enrichment folds with their 95% confidence intervals estimated by partitioned LDSC. Exclusively the 8 bordeaux modules are significantly enriched for common SCZ risk. **(H)** Number of genes mapping to SYNGO BP terms (top) and significantly enriched SYNGO terms (bottom) for bordeaux module genes shared between at least two of the 8 cell types.

### Significant enrichment for genetic schizophrenia risk exclusively in the bordeaux gene-program

To reveal which modules are enriched for common genetic schizophrenia risk, we performed a SNP-heritability analysis (Finucane et al. 2015). For this purpose, we combined the two groups of modules (e.g. bordeaux and midnight modules) due to their high similarity at the gene level (Fig. 5F). Strikingly, we found that the bordeaux module is the only module that was significantly enriched for the schizophrenia SNP-heritability (enrichment 1.48, p = 0.001; FDR = 0.037, Fig. 5G). Even after combining the 8 bordeaux modules together, they neither contain the most genes (237 unique genes), nor the highest proportion of SNPs in comparison to other modules or the other module group (Fig. S20A). Since the bordeaux module comprises genes involved in a broad range of pathway categories, we were interested in which of these pathway categories confer most of the genetic risk and performed a gene-level genetic enrichment for each bordeaux module gene using mBATcombo (A. Li et al. 2023). Significant genes were also mostly shared between all or large subsets of cell types of the bordeaux modules (Fig. S20B). On the functional level, the genetic risk for schizophrenia is mostly attributable to ion transport, energy (ATP), and gene expression pathways (Fig. S20C). Remarkably, all bordeaux modules contain several mitochondrial DNA and nuclear mitochondrial genes (Fig. S20D, S22B). While most nuclear mitochondrial genes showed down-regulation (Fig. 3A,C,G), most mitochondrial DNA genes were up-regulated in neurons (except excitatory layer 5/6 IT), OPCs and microglia. Bordeaux module genes shared between at least two of the 8 cell types were significantly enriched for pre- and post-synaptic SynGO pathways, including pre- and postsynaptic cytoskeleton and neuronal dense core vesicles (Fig. 5H).

### Energy deficiency, synaptic dysfunction and nervous system disruptions in SCZ samples confirmed by proteomics and long read RNA-seq

The bordeaux module gene enrichments are in agreement with proteomic analyses on these cases and controls (Table T1, Koopmans et al.) where mitochondrial, energy and synaptic pathways were enriched (Fig. S21A). Interestingly, mitochondrial genes were mostly unchanged in proteomic analyses (Fig. S22B), with the exception of MT-CO3. In total, 89 unique bordeaux module genes were also detected in proteomics. Protein abundance was increased by a log2 fold change of more than 0.5 for TUBA1B, MT-CO3, MT-ND3 and SLC1A2 and decreased by more than 0.5 for SNCG, VGF, and HPCAL1 (Fig. S21B). Proteomics (Koopmans et al.), long read RNA-seq (Abrantes et al.) and snRNA-seq (DEGs, network modules, and TWAS analyses) functionally overlap in nervous system and energy deficiency pathways (Fig. 6A). The latter were specifically attributable to the mitochondrial respiratory chain complexes I and IV of the mitochondrial inner membrane and the mitochondrial proton-transporting ATP synthase complex (Fig. 6B). RNA processing and binding, similar to Chromatin and DNA organization pathways, were observed in up-regulated DEGs, TWAS and long read RNA-seq (Fig. 6A,C). While ion transport gene expression was altered according to long read and snRNA-seq, synaptic pathways and cell-cell signaling were identified by proteomics, long-read and snRNA-seq. Interestingly, all three modalities identified extracellular matrix and microtubule pathways to be affected in SCZ (Fig. 6A-B).

**Figure 6.**
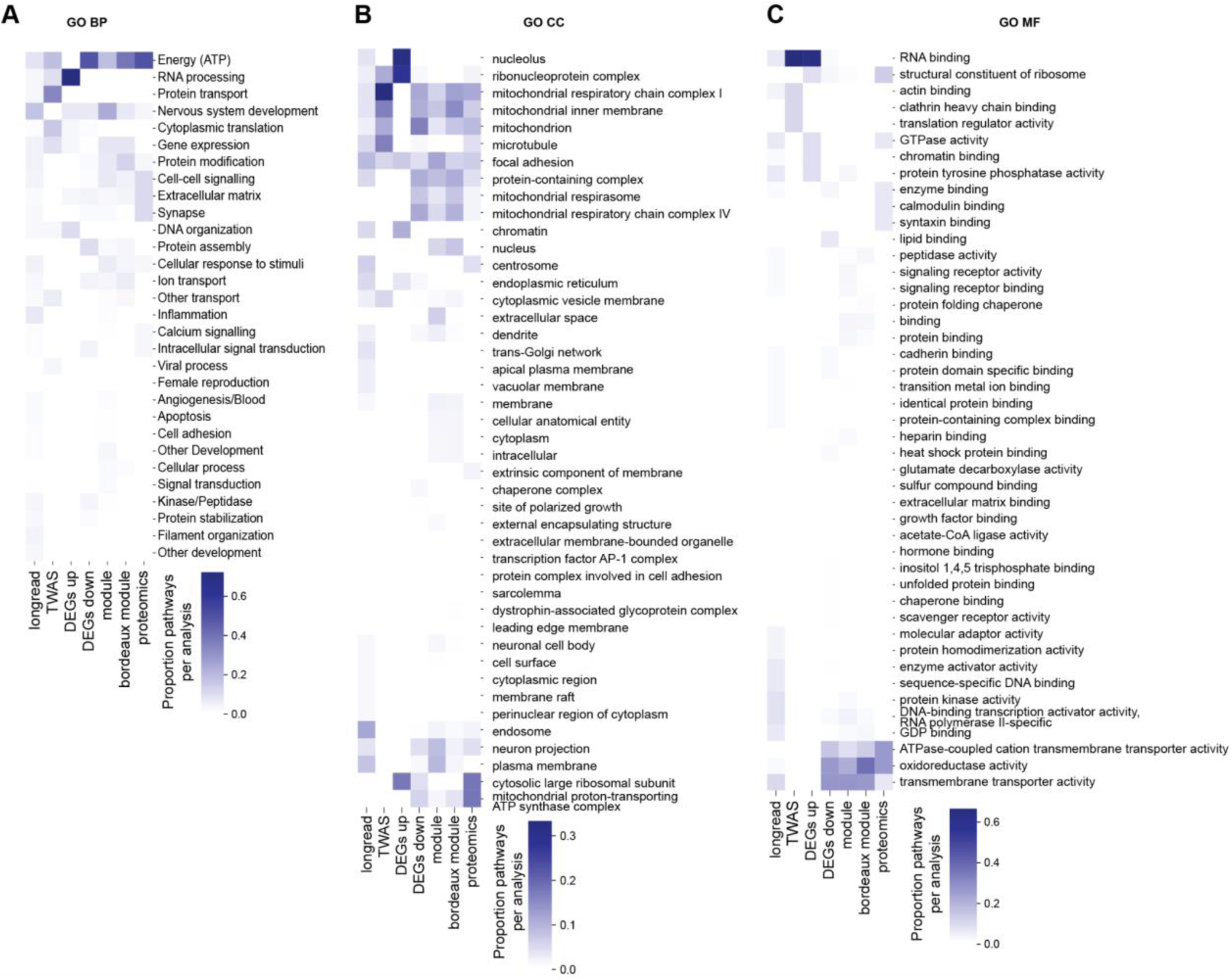
Proportion of **(A)** GO BP **(B)** cellular component (CC) and **(C)** molecular function (MF) pathways perturbed per analysis performed: DEG, GRN and TWAS analysis on single cell transcriptomics, DIUGs on long read RNA-seq, DAP coding genes on proteomics.

## Discussion

In this study, we genotyped and sequenced RNA in single nuclei of 43 human post-mortem tissue samples of the prefrontal cortex of schizophrenia patients and 42 age-, sex- and PMI-matched controls. With cell type resolution, we performed analyses across multiple levels, including cell type abundance, DEG, eQTL and TWAS, gene co-expression network inference and differential network analysis, each in combination with pathway enrichment analysis and common schizophrenia risk enrichment analysis. Our study reveals the transcriptomic perturbations in specific cell types and, by integration with genotyping data, distinguishes aetiological and downstream effects of the disorder. Thus, we provide a map of biological processes involved in the development of schizophrenia, and of downstream functional impairments in schizophrenia, both with cell type-resolution. Moreover, our analyses suggest a partial answer to the meaning of GWAS in schizophrenia (Patrick F. Sullivan, Yao, and Hjerling-Leffler 2024), that the common variant genetic basis of schizophrenia manifests partly via gene expression changes in *cis*.

We observed no significant differences in cell type abundances for our cohort. This finding is in agreement with another snRNA-seq study of similar scale (Ruzicka et al. 2024), but conflicts with the first report from a smaller snRNA-seq study (Batiuk et al. 2022). This finding is also noteworthy because we found diminished expression in schizophrenia of genes involved in cell differentiation. Together with other conflicting evidence for overall changes in neuronal numbers in the prefrontal cortex of schizophrenic patients (Pakkenberg, Scheel-Krüger, and Kristiansen 2009), this warrants careful interpretation of results from induced pluripotent stem cell experiments, where changes in progenitor proliferation and cell type composition have been reported (Notaras et al. 2021; Sawada et al. 2020; Stachowiak et al. 2017). Another caveat is our limited resolution in cell types (we analyze 16 and 37 cell classes/types while 100s have been suggested) which means that differences reported in smaller neuronal populations (subtypes) might have eluded our detection (Adorjan et al. 2020). It is also possible that cell type abundance changes are specific to a subset of individuals and associated to different comorbidities, medication profiles, etc.

In general, we find that genes implicated by common risk variants are down-regulated while rare variants are either unchanged or up-regulated, with the exception of CAGNA1G. This supports a model where common genetic risks drive the network towards a disease-state mainly through down-regulation of genes. We also confirmed that common genetic risk is mostly attributable to excitatory neurons (Skene et al. 2018, Yao et al 2024) and not in non-neuronal cells. In line with this both our DEG and heritability enrichment analyses point towards pronounced perturbations in excitatory neurons, with excitatory L2-3 IT neurons as the most affected cell type in schizophrenia. This latter finding confirms reports from two other snRNA-seq studies (Ruzicka et al. 2024; Batiuk et al. 2022).

Our gene co-expression network analysis also implicated a schizophrenia-relevant network of genes in layer 2-3 IT II cells which was related to peptidergic signaling and developmental processes. Core genes in the module included glutamatergic synapse genes *HOMER1* and *NPTX2* which were confirmed as decreased in the proteomic analysis (Koopmans et al.). This module did not reach the statistical threshold for heritability enrichment but was the second most enriched network in the list, suggesting it might still be of aetiological relevance. Although we find most DEGs in excitatory layer 2-3 IT neurons, we also found deep layer neurons to be perturbed in schizophrenia, and that these DEGs confer a substantial amount of the genetic risk for schizophrenia, perhaps because many of the genes were synaptic in comparison to lists of DEGs in other neuronal cell types.

The diversity of GO pathways enriched in down-regulated genes across cell types suggests a heterogeneity in the mechanism of schizophrenia. Our DEG results highlighted the disturbance in mitochondrial and ATP metabolism terms in the down-regulated genes both in neurons and astrocytes, suggesting a broad impact across multiple cell types. This finding is supported by the first smaller snRNA-seq study (Batiuk et al. 2022) but was not reported in two larger snRNA-seq schizophrenia control studies (Ruzicka et al. 2024, Ling et al., 2024). This is puzzling, but could possibly be explained by the common use of mitochondrial genes as a quality metric to filter cells/nuclei in sc/snRNA-seq, since Ruzicka et al. 2024 excluded nuclei with more than 10% UMIs in mitochondrial DNA genes from their analysis while (Batiuk et al. 2022) and our study included these genes in the analysis. We observed that mitochondrial DNA genes were up-regulated in several neuronal types, OPCs and Microglia. Increased mitochondrial DNA gene expression has previously been observed in major depression disorder and stress mouse models (Cai et al. 2015) Mitochondrial dysfunction has also been reported in a study analysing organoids derived from bipolar disorder patients (Phalnikar et al. 2024). That our findings were consistent with results from the accompanying schizophrenia study using proteomics of a largely overlapping cohort (Koopmans et al.) is encouraging since it greatly negates the risk of mitochondrial dysfunction being a false finding related to single-nuclei isolation and highlights the strengths of a multimodal approach. Additionally, in both studies, transcriptomic and proteomic, mitochondrial inner membrane components were more affected than outer membrane components suggesting pathway specificity in regulation of mitochondrial function. Gene network analysis identified modules across several cell types containing genes related to mitochondrial function which were significantly enriched for schizophrenia heritability, and TWAS analysis also pointed to the respiratory complex in neuronal cell types. These changes are thus likely causal rather than a response to disorder-related changes in the network. Mitochondrial dysfunction has been implicated with schizophrenia before (Roberts 2021; “Mitochondrial Dysfunction in Schizophrenia: Pathways, Mechanisms and Implications” 2015)Rajasekaran et al. 2015), is important for adolescent brain development (Ene et al. 2023), and synaptic function via cytoskeletal mechanisms (Marchisella, Coffey, and Hollos 2016). Moreover, schizophrenia related RNA hypoediting preferably happens in mitochondrial genes (Choudhury et al.).

For up-regulated genes we observed increased expression of genes involved in RNA processing across both excitatory and inhibitory neurons. This list included RNA-binding proteins and core genes in the nuclear speckles where a majority of mRNA splicing occurs. Long-read sequencing analysis confirmed that not only are these genes more expressed but they themselves undergo alternative splicing in patients (Abrantes et al.). Up-regulation of proteins in these categories was not seen, which could perhaps be attributable to differences in detection sensitivity and/or because many of the nuclear speckles genes are structurally long non-coding RNA genes. In addition, our TWAS analysis was also significantly enriched for the RNA-binding pathway. Still, the enrichment of schizophrenia heritability was not significant in the DEG-derived RNA-binding pathways which leaves an aetiological role for this pathway an open question. In support of this RNA-binding proteins, which regulate mRNA transport and protein translation at synapses (Sephton and Yu 2015), have previously been implicated in schizophrenia genetic risk (Velázquez-Cruz et al. 2021; Park et al. 2021).

Compared to other non-neuronal cell types, astrocytes had the highest similarity with neuronal cell types in the biological processes in down-regulated genes including synaptic specific ontology terms. This is in alignment with (Ling et al. 2024) who recently identified a synaptic neuron and astrocyte program using latent factor analysis. Since astrocytes contribute to homeostasis of neurotransmitters and energy in neurons and this interaction underlies extracellular ion homeostasis, volume regulation, and neuroprotection in the central nervous system (Benarroch 2005), our findings lend support to a perturbed interaction between neurons and astrocytes in schizophrenia.

Working with postmortem data introduces important limitations. We were technically limited to sequencing isolated nuclei instead of whole cells, so our results may not faithfully reflect the total RNA content in the cell (Skene et al., 2018). Postmortem data also mean single time point measurements, which complicates identifying causal relationships and disorder dynamics. Differences in cell type abundances creates large differences in the power of analysis between common and rare cell types. This study mostly contained subjects of European ancestry due to tissue sample availability. As we saw examples of mismatches between stated and measured ancestry, this highlights the usefulness of genetic analysis of the samples. The sample size could always be larger, and our findings require replication. Importantly, some of the effects observed in patients may have resulted from medication rather than the disorder. Toxicology reports were available but they only reflect very recent medication use and not long-term medication history. Measuring the effect of medication on brain cell transcription is not an easy task but studies in primate models (Martin et al. 2015) and comparing drug naive and medicated patients (Schulmann et al. 2023) suggest that the main effects of antipsychotics partly seem to counteract disease-related changes, which if anything should mainly decrease the disease signal detected. Of course, schizophrenia patients are regularly prescribed additional medications beyond antipsychotics, including antidepressants and mood stabilizers which potentially introduce additional variability. Inclusion of clinical data, such as extensive medical history and detailed phenotypic characterization, might enhance future efforts. Finally, our study focused on prefrontal cortical regions, but additional brain regions, including hippocampus and amygdala, are also enriched for schizophrenia heritability. Identifying cell-type specific transcriptional changes in these regions will likely move us closer towards an understanding of the disease-biology of schizophrenia.

In this study, we performed single-nucleus RNA sequencing and genotyping of post-mortem prefrontal cortex tissue from schizophrenia patients and matched controls, providing a cell type-specific map of transcriptomic alterations. Our findings, also largely supported by multi omic analysis, indicate that common schizophrenia risk variants are predominantly associated with gene down-regulation, especially in excitatory neurons, while rare variants show an up-regulation pattern, highlighting a potential difference in disease mechanism. We observed significant transcriptomic perturbations in excitatory layer 2-3 neurons, with mitochondrial dysfunction and RNA processing alterations implicated across multiple cell types. A synaptic phenotype was present in deep layer excitatory cells and astrocytes. These results deepen our understanding of the cell type-specific molecular underpinnings of schizophrenia, while technical limitations and potential medication effects call for further investigation to disentangle these findings. Future studies, on a larger scale and a more globally representative sample, are needed to better understand patient-level heterogeneity and might help to reconcile some of the disparate findings from different data sets.

## Methods

### Subjects

We obtained human Brodmann Area 8/9 tissue from 43 schizophrenia subjects and 42 control individuals from three sources, NBB (Netherlands Brain Bank), Craig A. Stockmeier (University of Mississippi Medical Center), Macedonian/New York State Psychiatric Institute Brain Collection (Andrew J. Dwork, Columbia University). All subjects included in the study were matched between the two groups for age, gender, and postmortem interval (PMI, Fig. 1A, Table T1).

Each case was assigned a consensus diagnosis based on a review of medical records and a questionnaire completed by the donor’s family. Demographic variables for the assembled cohort are listed in Table T1. Upon arrival at Karolinska Institutet, the tissue was stored at −80°C prior to usage.

### Sample preparation

After verifying that sampled tissue block included all six cortical layers and underlying white matter, frozen tissue was cut, thawed, ground and homogenized in batches of 8 samples per day (four schizophrenia and four sex-, age- and PMI-matched controls). Tissue homogenates were filtered twice and pelleted by centrifugation to isolate nuclei. To enrich 85% of neurons, nuclei suspension was labeled with NeuN antibody and sorted by FACS (Supplemental information 2). For a few samples, pairs were, due to technical reasons, distributed across different sequencing runs.

### Single-nuclei RNA sequencing

The pooled nuclei were then applied to one of eight channels of one 10x Genomics Chip B, targeting recovery of 5,000 nuclei per channel. As the Chromium system only allows for a maximum of eight samples per capture run, multiple batches were required to collect the nuclei for all 86 samples (18 batches). 10x Genomics libraries were prepared as per standard protocol (10x Genomics Chromium Single Cell protocol, CG000183_ChromiumSingleCell3_v3_UG_Rev-A.pdf). All snRNA-Seq libraries were sequenced, in five batches, on an Illumina NovaSeq instrument (18-20 libraries per NovaSeq S2 flowcell) to achieve an average sequencing depth of 120,000 reads per cell. Gene count matrices were generated by aligning reads (including intronic reads) using 10x Genomics Cell Ranger software v6.0.1.

### Quality control of samples and cells

To filter low quality nuclei, we removed outliers with pagoda (Barkas et al. 2021, Fig. S1A,D), applied threshold (Fig. S1B,D), and doublet (Wolock, Lopez, and Klein 2019, Fig. S1C,D) filtering (Supplemental Information 2). Subsequently, we removed samples from the data set that contained less than 500 nuclei after those filtering steps. Additionally, we performed principal component analysis on a range of quality control metrics (Table T2) to identify outlier samples (Fig.S1 E,F).

### Data integration, clustering, and cell type annotation

We integrated donor samples (Fig. S2A), and clustered the integrated data by using conos (Barkas et al. 2019, Petukhov et al. 2021, Fig. S2B). These clusters were subsequently informed by automated cell type annotations of their nuclei (Fig. S2C). To automatically annotate a given nuclei with the most probable cell type out of 76 different cell types from a reference data set (Hodge et al. 2019), we utilized scmap (Kiselev, Yiu, and Hemberg 2018) with a similarity score of at least 0.5. According to these annotations, we divided cells in three main classes: non-neuronal cells, excitatory neurons and inhibitory neurons (Table T3) and subsequently performed a second level clustering and annotation for each class separately (Fig. S3, Table T4, Supplemental Information 4). We confirmed the automatic annotations by visualizing the expression of established marker genes (Fig. S4A). As expected, non-neuronal cell types showed a lower number of genes expressed compared to neurons (Fig. S4B). Cell types showed similar distribution across donor samples and between the groups (schizophrenia and control, Fig. S4C).

### Cell type abundance analysis

We utilized the cluster-based approach in cacoa (Petukhov et al. 2022) with default settings to test if the cell type clusters obtain a higher or lower abundance in schizophrenia as compared to the control group samples (Fig. S1F).

### Differential gene expression analysis

Differential gene expression (Fig. 2B-E, S6, Table T5) was performed using DESeq2 (Love et al. 2014) on cell type-specific pseudo bulk data. Genes on X- and Y-chromosomes and with low counts (at least 70% of samples with more than 10 counts) were removed. We performed DESeq2 for each cell type separately, which includes normalization with DESeq2’s implemented size factor estimation function. We performed removal of unwanted variation with residuals (RUVs) analysis (Risso et al. 2015, Fig. S5, Supplemental Information 5), in order to control for technical variation and batch effects in the pseudobulk counts. We included the estimated RUV factors in the full and reduced models that we tested with the likelihood ratio test in DESeq2. For differential testing, the alpha value was set to 0.05 and the FDR was controlled with the Benjamini-Hochberg procedure (Benjamini and Hochberg 1995).

### Pathway enrichment analysis

In order to test the enrichment of ontology gene sets for identified DEGs (Fig. 3, S8-10, Table T6), we performed Gene Set Analysis (GSA) using the hypergeometric test. We ran GSA separately for up- and down-regulated genes (adjusted p-value<=0.3) of each of the 16 cell types. As the background gene list we chose 13626 genes known to be expressed in the brain cortex. For a pathway to be tested, we requested an overlap of at least 3 up- or down-regulated genes, respectively. Multiple testing correction was conducted using Benjamini-Hochberg procedure (Benjamini and Hochberg 1995) within each subgroup (e.g. GO BP, Table T6). To group significant gene ontology terms into categories, we used rrvgo (Sayols 2023).

### Gene co-expression network inference

GRN inference was performed using high dimensional weighted gene co-expression network analysis (hdWGCNA, Morabito et al. 2023) on the 2000 most variable genes of each cell type. For metacell calculation of snRNAseq data we required a gene to be expressed by at least 5% of nuclei and at least 250 nuclei in a particular grouping (cell type and donor). We calculated meta cells using 50 nearest neighbors, and allowed for not more than 13 nuclei being shared between each two meta cells (Supplemental Methods, Fig. S19A-C). Excitatory layer 5-6 IT neurons II, microglia, endothelial and mural cell clusters were removed from the network analysis, since they either had too few cells and/ or donors to construct the metacells or the network reliably. Using the metacell data, we constructed cell type specific networks (Fig. 5A,C) on the 2000 most highly variable genes of each cell type and tested for each module differential expression in schizophrenia using the likelihood ratio test (‘LR’ setting in hdWGCNA, Fig. 5B). For all significant modules we tested module genes for enrichment of GO pathways by using gprofiler2 (Fig. 5D, S19D, Table T8). To group significant gene ontology terms into categories, we used rrvgo and merged the parent categories manually to reduce the number of categories further for visualization purposes (Fig. 5D, Table T9).

### Heritability enrichment analysis

We primarily focused on SNP-level heritability to evaluate the enrichment of schizophrenia genetic risk in selected gene lists using partitioned LDscore regression (with the 1000 Genome European reference panel, (Finucane et al. 2018). For cell-type-specific gene lists, we tested 1) specifically expressed genes per cell type and 2) top down- and up-regulated genes per cell type. For gene lists not specific to cell types, we tested 3) GO terms suggested by DEGs and 4) in hdWGCNA modules. We also performed gene-level analysis evaluating the mBATcombo-significant genes in 3) and 4) using hypergeometric tests (detailed in supplementary Method).

### Genotyping, genetic risk score, and CNV calling

Genotyping was performed using the GSAv3 Illumina ChIP in 2 batches. Genotype quality control (QC) was performed in Ricopili (Lam et al. 2020), and imputation was performed according to the Haplotype Reference Consortium reference panel (HRC.r1-1, Lam et al. 2020; McCarthy et al. 2016) in GRCh37. We removed SNPs with minor allele frequency (MAF)<0.005 and INFO<0.1 per batch, merged the genotyping batches and derived the principal components (PCs) which were used as covariates for the CNV calling and eQTL analysis. We then derived the polygenic risk score (PRS) of schizophrenia using SNPs with GWAS p-value≤0.1 (Trubetskoy et al. 2022) and clumped at r^2^<0.1 in 1 Mb windows; SNPs with missingness> 5%, INFO<0.4, or MAF<0.05 and individuals (n=2) with >2% missing genotypes were excluded. PRS were calculated as the sum of dosages of the post-clumping SNPs weighted by the effect size from the GWAS (PLINK v1.9). The extended MHC region (chr6:25-34 mb) was removed.

Copy number variants (CNVs) were called for autosomes and chromosome X (Szatkiewicz et al. 2014; Farrell et al. 2023) using EnsembleCNV which packages three CNV calling algorithms (Zhang et al. 2019). Per algorithm and per genotyping batch, segments with <10 probes were removed, and segments with a gap <20% of the segments were joined. We further removed CNVs <100kb and CNVs with over 50% overlap with large genomic gaps, rearrangement segment, and genomic super duplications (Farrell et al. 2023). We then kept CNVs detected by at least two algorithms with ≥50% reciprocal overlap and used the outer boundary from all algorithms as the final boundary of the CNV. No sample was excluded based on the first 3 PCs of sample statistics (LRR_SD, BAF_SA, WF, number of CNVs from the three calling algorithms) with 6 standard deviations as the threshold. Next, a CNV burden test was performed to compare the differences in the number and size of CNVs between cases and controls, adjusting for the first 5 genotyping PCs and genotyping batch. We also explored whether the identified CNVs had ≥50% reciprocal overlap with known schizophrenia-relevant CNVs (detailed in Supplementary Table 12, (Singh et al. 2022)).

### eQTL analysis

eQTL analysis was performed using data from 73 samples with European ancestry, using QTLtools (version 1.3.1, Delaneau et al. 2017; Y. E. Li et al. 2023). To avoid overfitting, we limited the number of covariates and adjusted for phenotype status, genotyping batch, the first 2 genotyping PCs, and the first 4 expression PCs in the corresponding cell class. Gene expression was aggregated across cells per donor per cell class or type and normalized to CPM. For each included gene, we considered SNPs within the 1Mb window around the transcription starting site (TSS, based on GENCODE v40, 4/2021). We calculated q-value (FDR corrected nominal p-values) to account for multiple testing per cell class/type, and SNP-gene pairs with q-value≤0.05 were considered significant eQTL pairs. We further evaluated the agreement between our eQTL effects and previously published bulk and cell type eQTLs and the agreement of our eQTL effects between cell classes. Finally, we examined the overlap between our eQTLs and predicted cell type specific functional genomic regions as suggested by candidate cis-regulatory elements (cCREs, (Delaneau et al. 2017; Y. E. Li et al. 2023); more details in Supplementary Method). All analyses were performed under genome build GRCh37.

### TWAS analysis

We integrated the cell class and cell type eQTLs with the GWAS summary statistics of schizophrenia (Trubetskoy et al. 2022) to investigate expression-trait association using CommS4 (Yang, Yeung, and Liu 2021). Next, we examined the enrichment of gene ontology pathways in the genes with TWAS FDR≤0.3 per cell type using hypergeometric test and grouped significant gene ontology terms (FDR≤0.05) into categories using rrvgo (Sayols 2023).

### Cross-modality/ -analyses pathway analysis

We pooled together pathway enrichment results of TWAS (any cell class), DEG (any cell type up-regulated genes and any cell type down-regulated genes), all module genes, bordeaux module genes, differential isoform usage genes detected by long read RNA-seq (Abrantes et al.) detected in a subset of our cohort, and differentially abundant proteins’ coding genes detected by proteomics of the same and a few additional samples (Table T1). All significant gene ontology pathways, identified with GSA based on the hypergeometric test, were re-structured using rrvgo (Sayols 2023) and for biological pathways categories were further merged to achieve lower resolution. For each modality and GO class (BP, CC, MF) we then calculated the percentage of significant pathways to observe commonalities and differences across modalities and analyses.

This study is approved by etikprövningsmyndigheten/ Swedish Ethical Review Authority (Dnr:2016/589-31).

## Supplemental Figures

**Figure S1.**
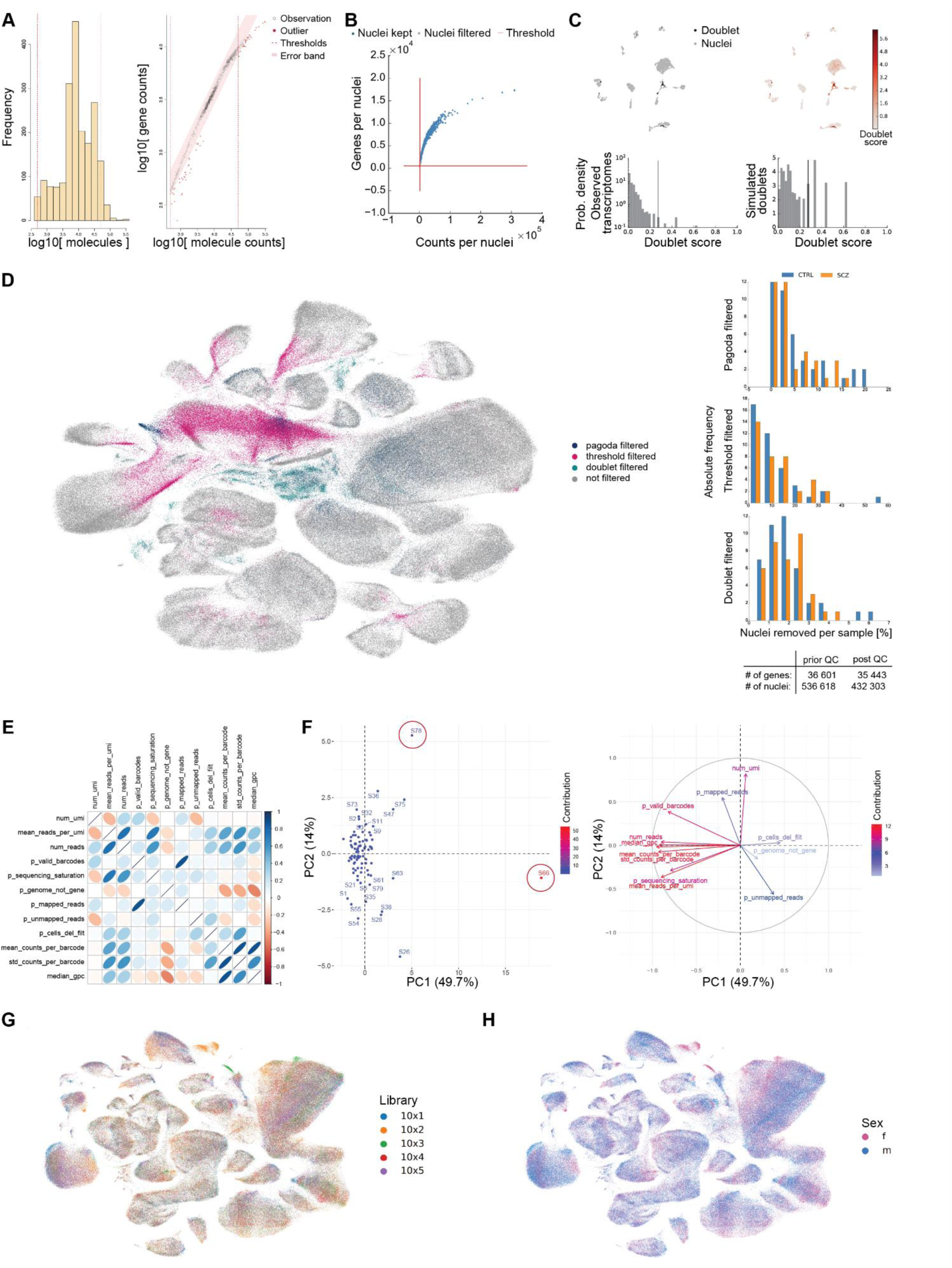
**(A)** Preprocessing of sample S1 (CTRL group) with pagoda indicating thresholds (red dashed lines) in histogram (left) and nuclei removed (red dots) and kept (black dots, right). **(B)** Counts vs. genes per nuclei of sample S1 indicating thresholds (red lines) applied to filter nuclei (gray dots). **(C)** Nuclei of sample S1 with their predicted doublet status (top left) and doublet score (top right) highlighted in UMAP downprojection. Histograms indicating observed (bottom left) and predicted doublet score distribution (bottom right). **(D)** First two dimensions of UMAP downprojection containing all 536 618 sequenced nuclei highlighting their filtering status (left). Histograms indicating percentage of nuclei removed per sample in each filtering step for schizophrenia (orange) and control group (blue) samples (right). **(E)** Correlation of each pair of 13 quality control (QC) metrics extracted per sample from snRNA-seq data. **(F)** First and second principal component (PC) resulting from PCA of weakly correlating QC metrics exposes two outlier samples S66 and S78 (left) and contribution of QC metrics to PC1 and PC2 (right). **(G)** UMAP highlighting cells in five different libraries. **(H)** UMAP highlighting cells from female (f) and male (m) donors.

**Figure S2.**
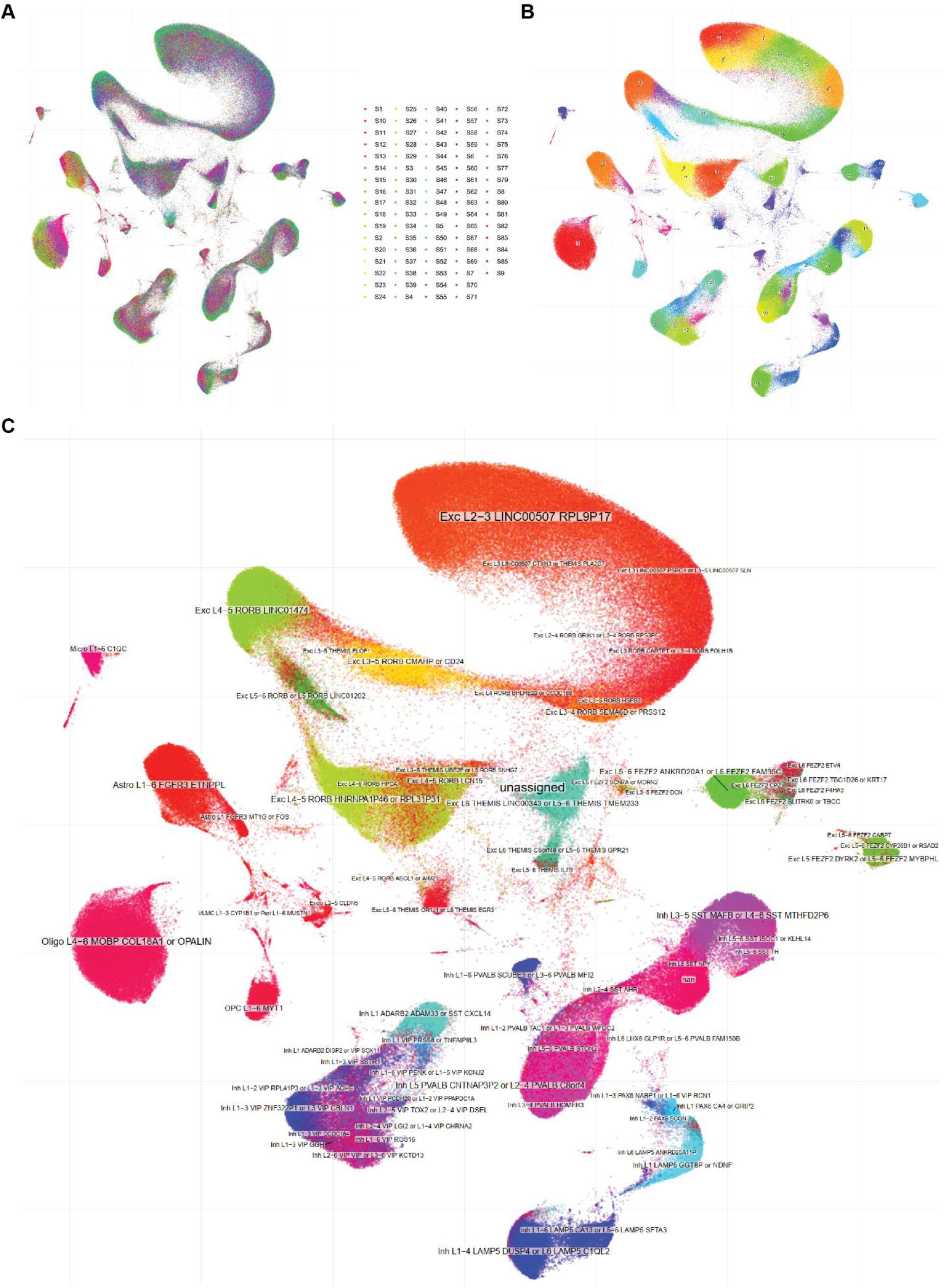
UMAP after data integration and clustering with conos highlighting **(A)** donor samples, (**B)** cluster IDs, and **(C)** cell type annotations.

**Figure S3.**
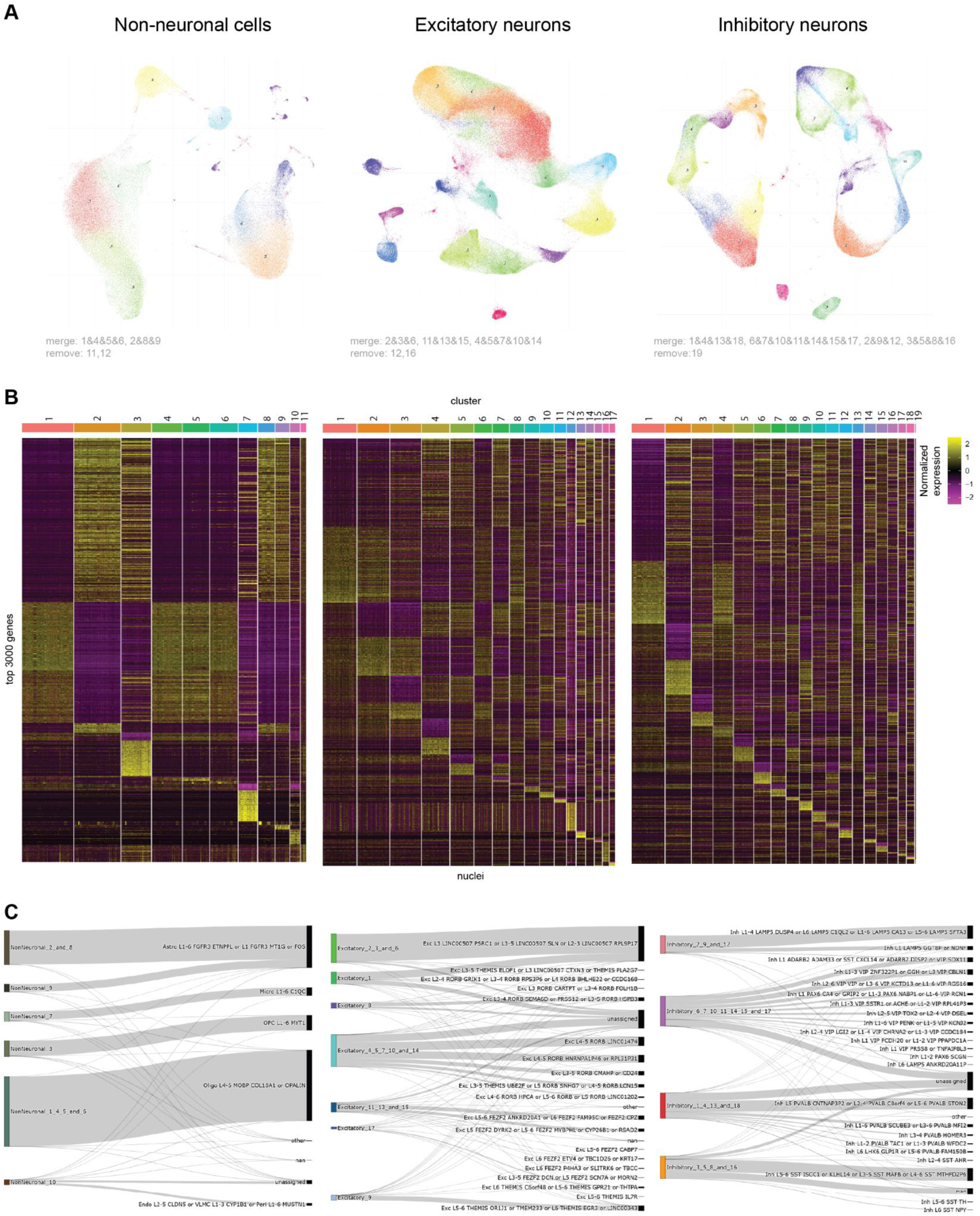
**(A)** UMAP after data integration and clustering with conos for Non-neuronal nuclei (left), excitatory (middle) and inhibitory neurons (right). (**B)** Heatmap of top 3 000 highly variable genes for each cluster in each cell type class (as shown in A). **(C)** Sankey plot indicating assigned proportions of cells in each cluster to cell types in reference data set. Nuclei got labeled as unassigned if their agreement score with any cell in the reference was below 0.5, unannotated nuclei in the reference are indicated as nan, and nuclei with label ‘other’ got a celltype from another class assigned.

**Figure S4.**
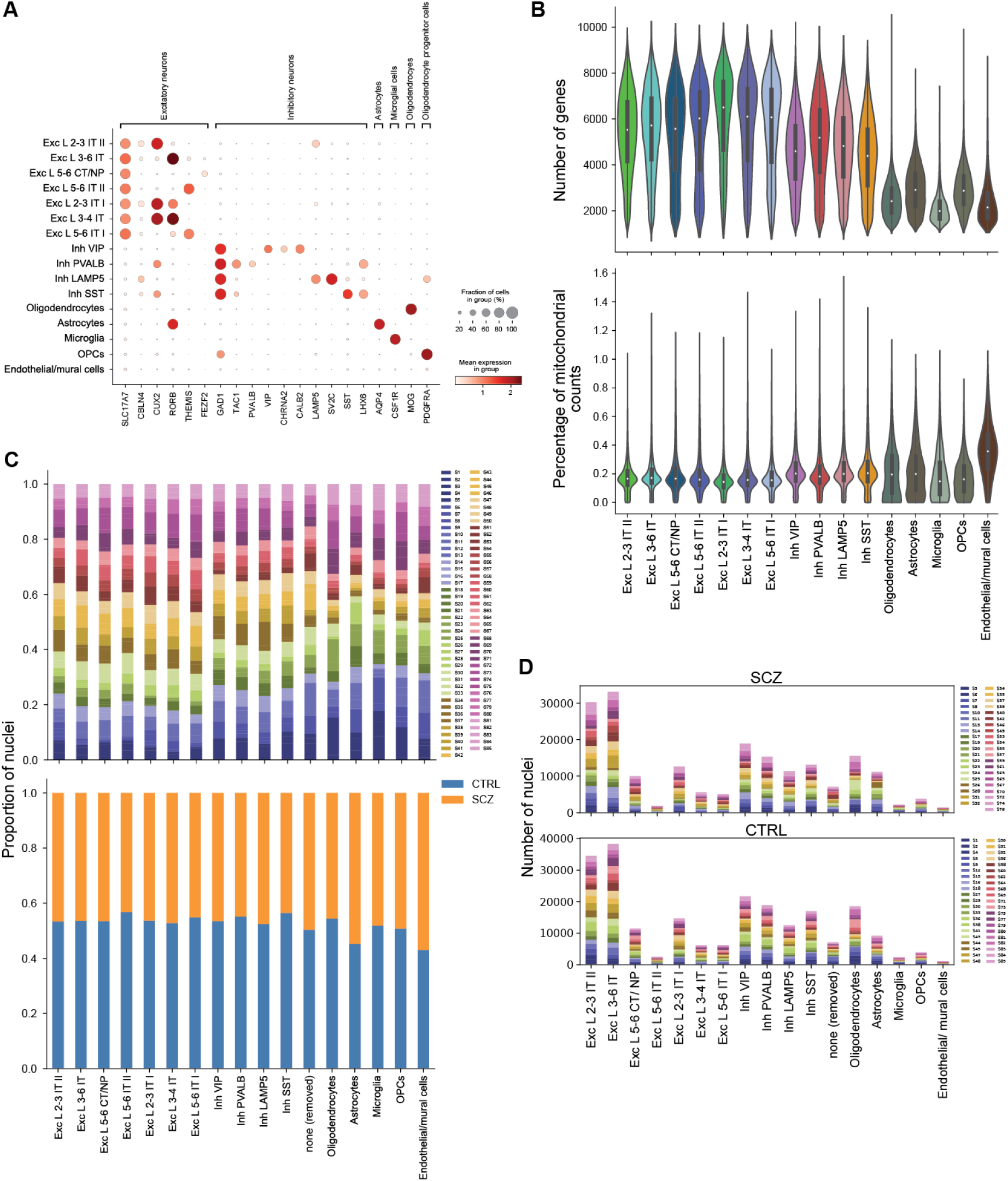
**(A)** Mean expression for known marker genes per cell type confirms annotation. Size of circles indicate percentage of cells of a cell type expressing the respective gene. (**B)** Distribution of number of genes (top) and of percentage of mitochondrial counts (bottom) per cell type. **(C)** Proportion of nuclei per sample (top) and group (bottom) for each cell type. **(D)** Number of nuclei per cell type for SCZ (top) and CTRL (bottom) samples.

**Figure S5.**
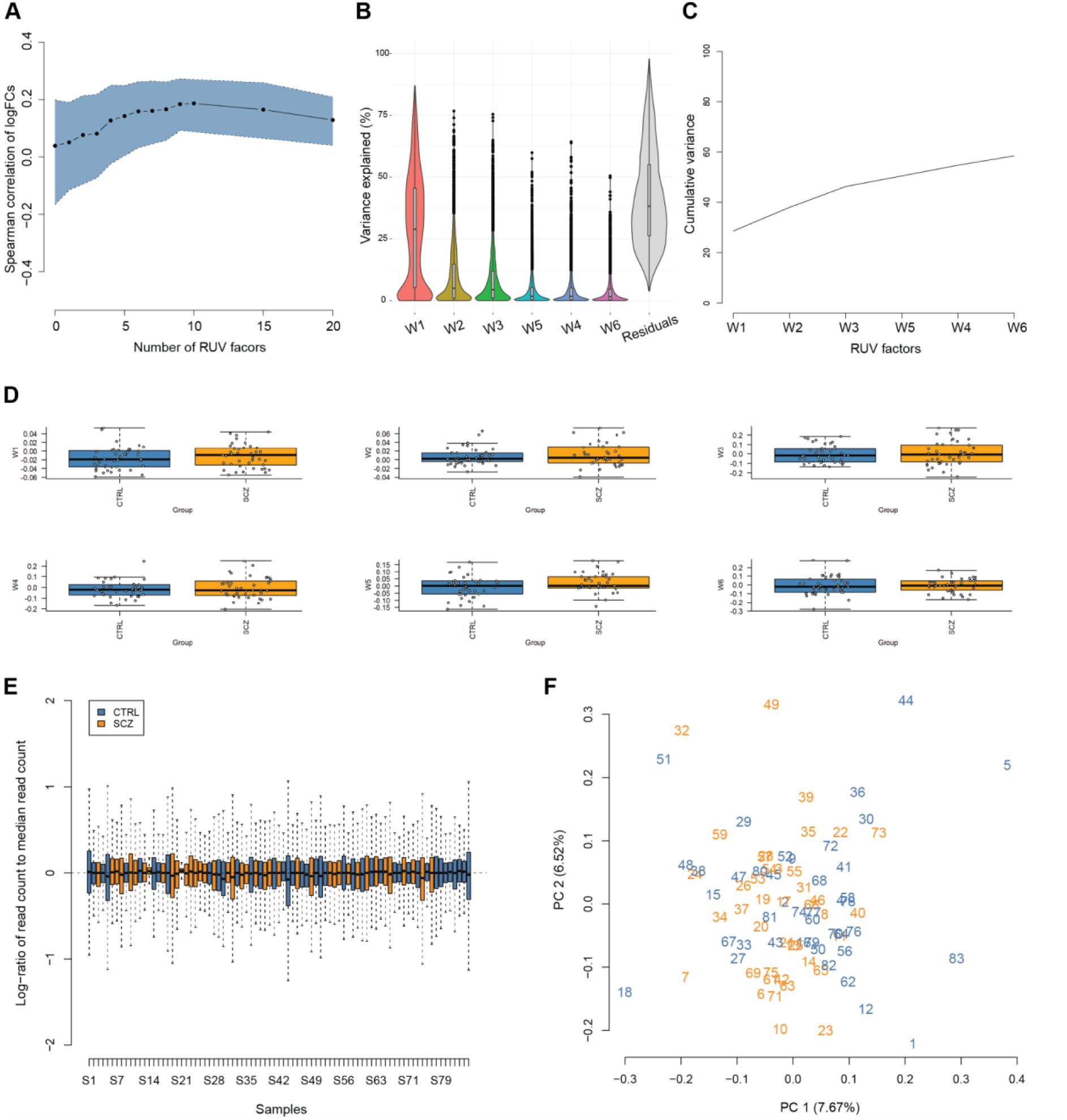
**(A)** Spearman correlation of log2 fold changes of both data halves maximized for 10 RUV factors. To avoid overfitting only six factors were used. (**B)** Explained variance distributions for the first six RUV factors and residuals. (**C)** Mean cumulative variance for the first six RUV factors, showing they explain together about 60% of the variance. **(D)** Box plots per group control (CTRL, blue) and schizophrenia (SCZ, orange) for each of the six factors. Center lines of box plots correspond to median, box limits to upper and lower quartiles, and whiskers to 1.5x interquartile range. Dots correspond to individual values.(**E)** Log-ratio of read count to median read count distribution per sample after RUV correction with six factors showing similar distributions. Center lines of box plots correspond to median, box limits to upper and lower quartiles, and whiskers to 1.5x interquartile range. (**F)** Principal component analysis of RUV normalized data with highlighted group status (schizophrenia samples in orange and control group samples in blue.

**Figure S6.**
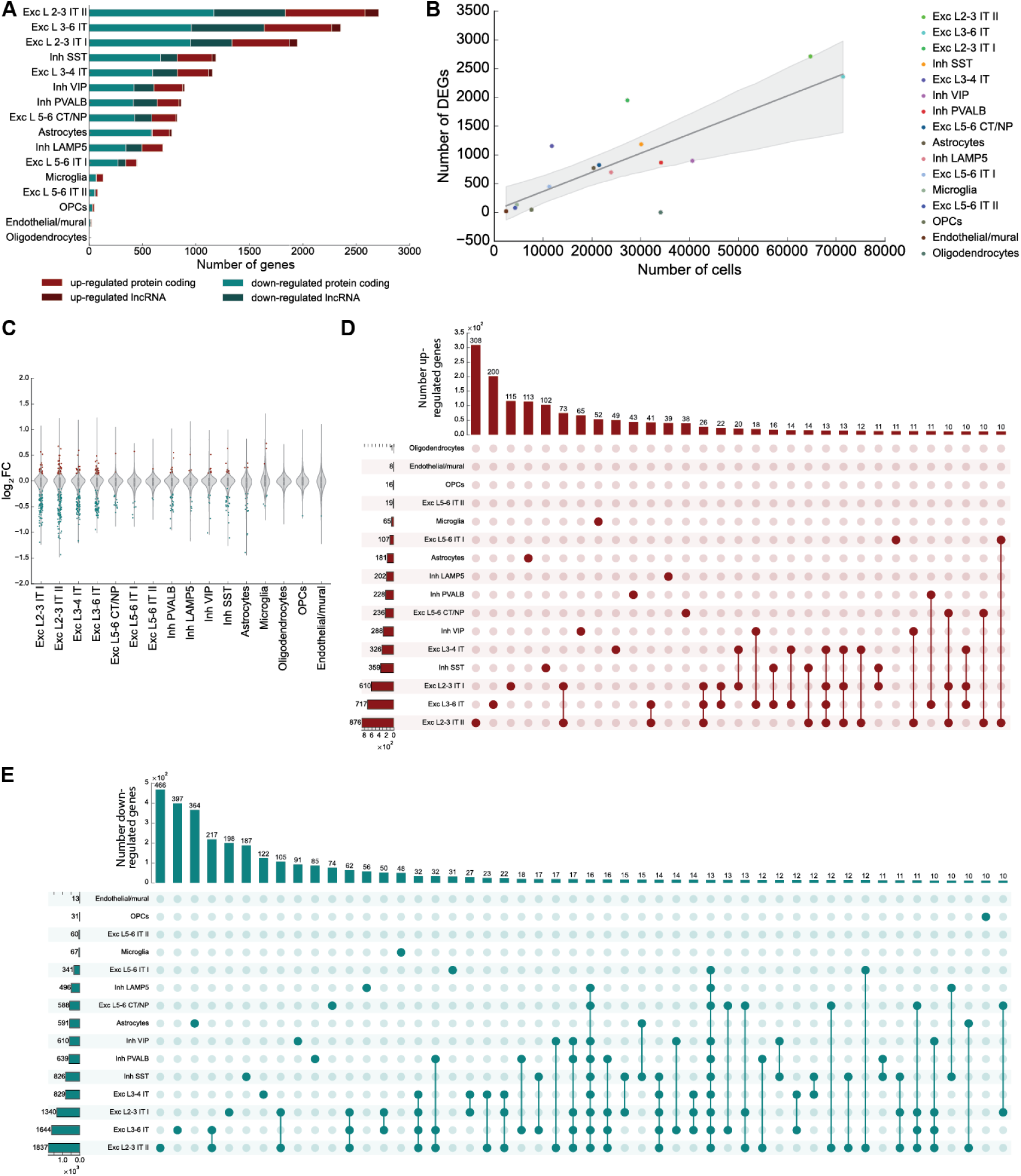
**(A)** Number of up- and down-regulated genes in schizophrenia identified per cell type cluster using threshold alpha=0.3. **(B)** Number of cells in cluster per number of identified DEGs shows also smaller cell type clusters ranking in top 3 regarding their number of DEGs for alpha=0.3. **(C)** Significantly up- (green dots) and down-regulated (red dots) DEGs obtain high absolute log2 Fold Changes. **(D)** Overlap of up- (top) and **(E)** down-regulated (bottom) genes in schizophrenia for alpha=0.3 between cell type clusters shows most DEGs are not shared between cell types and otherwise mostly shared between neuronal cell types.

**Figure S7.**
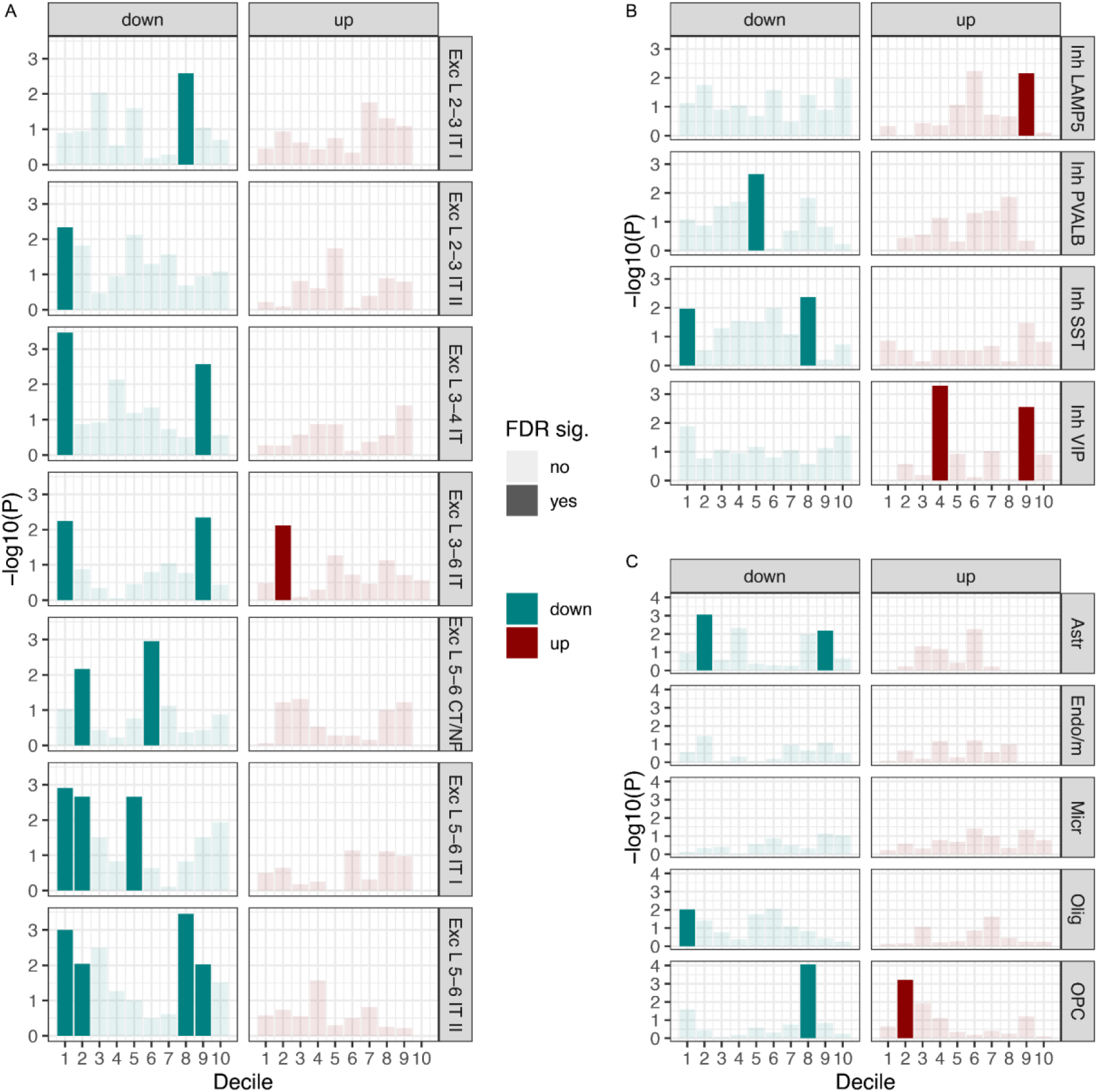
Enrichment of schizophrenia common genetic risk in deciles of DEGs in excitatory (A), inhibitory (B) and non-neuronal (C) cell types. The deciles were defined based on the adjusted p-value from DEG analysis; deciles 1-10 reflect higher to lower significance. Deciles in down-regulated genes are more likely to enrich common genetic variants of schizophrenia than deciles in up-regulated genes in general.

**Figure S8.**
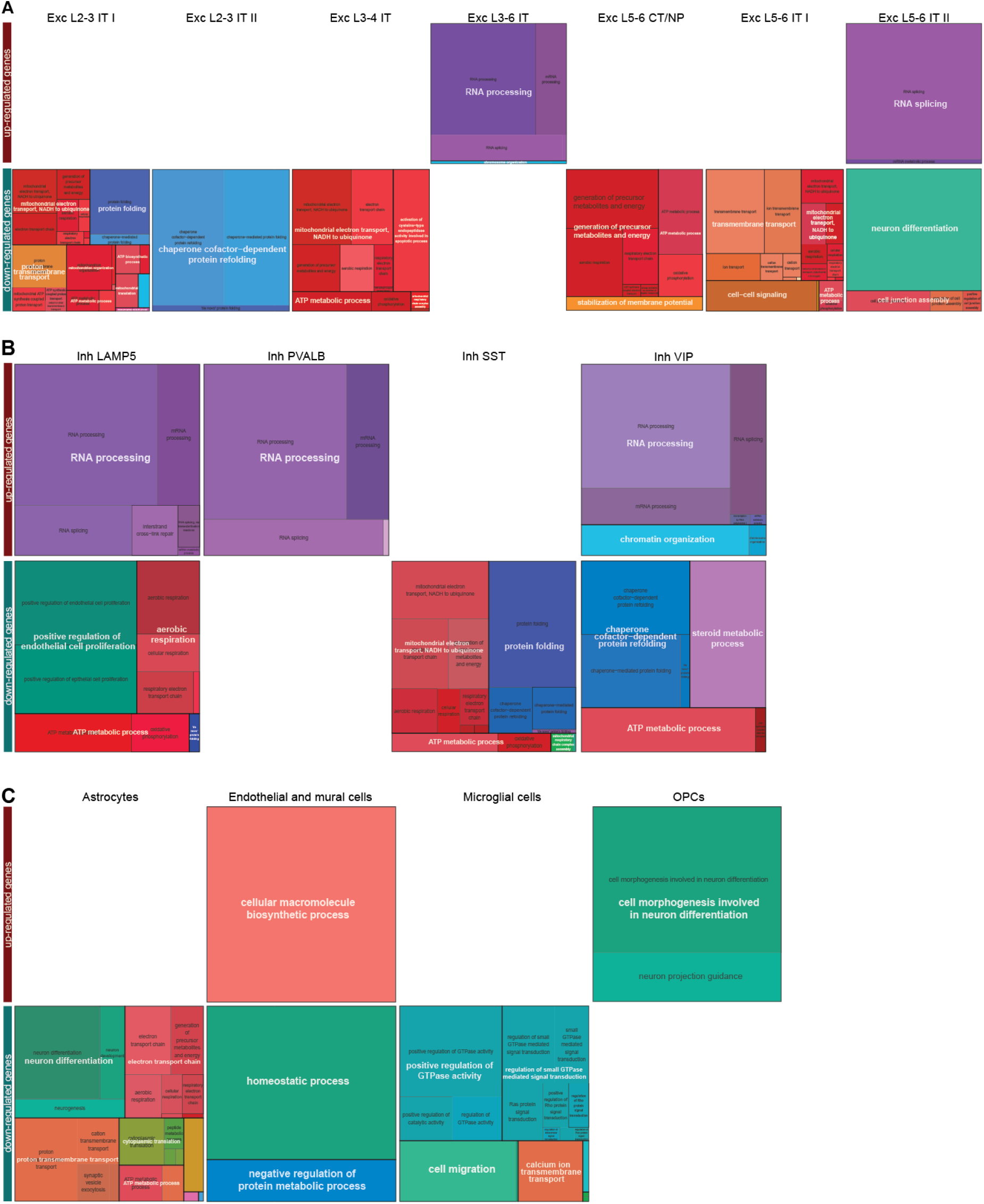
GO BP pathway categories for up- (top) and down-regulated (bottom) DEGs for each **(A)** excitatory **(B)** inhibitory neuron and **(C)** non-neuronal cell type.

**Figure S9.**
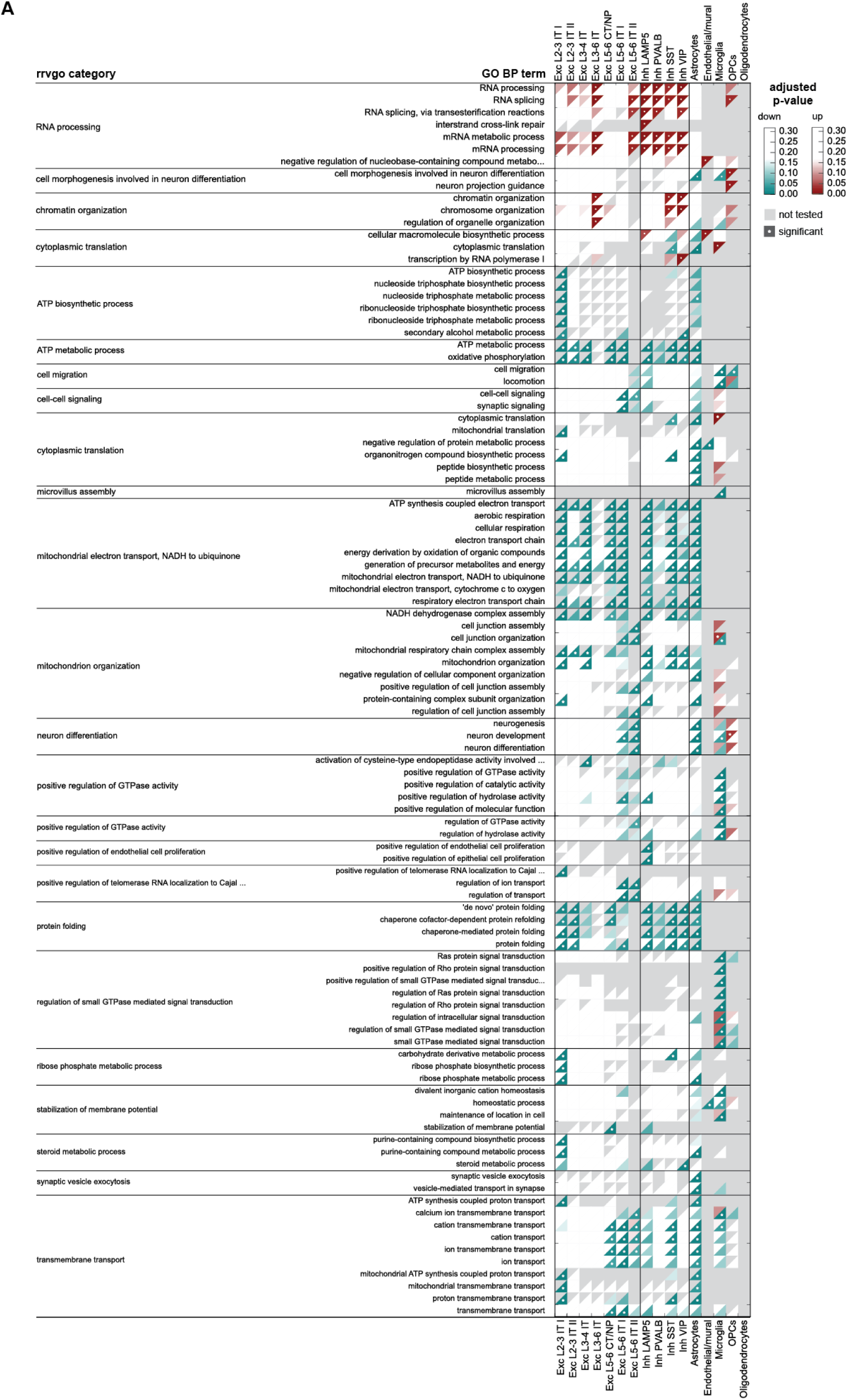
**A** GO biological processes (BP) enriched for up- (red) and down-regulated (green) DEGs. White dots indicate significant enrichment (adjusted p-value below 0.05) and gray triangles imply GSA analysis was not conducted due to insufficient overlap of DEG list with GO BP term. In total 1728 (12.7%) unique DEGs (alpha=0.3) were not mapping to any GO BP pathway and 796 (5.8%) unique DEGs (alpha=0.3) were not mapping to any GO pathway.

**Figure S10.**
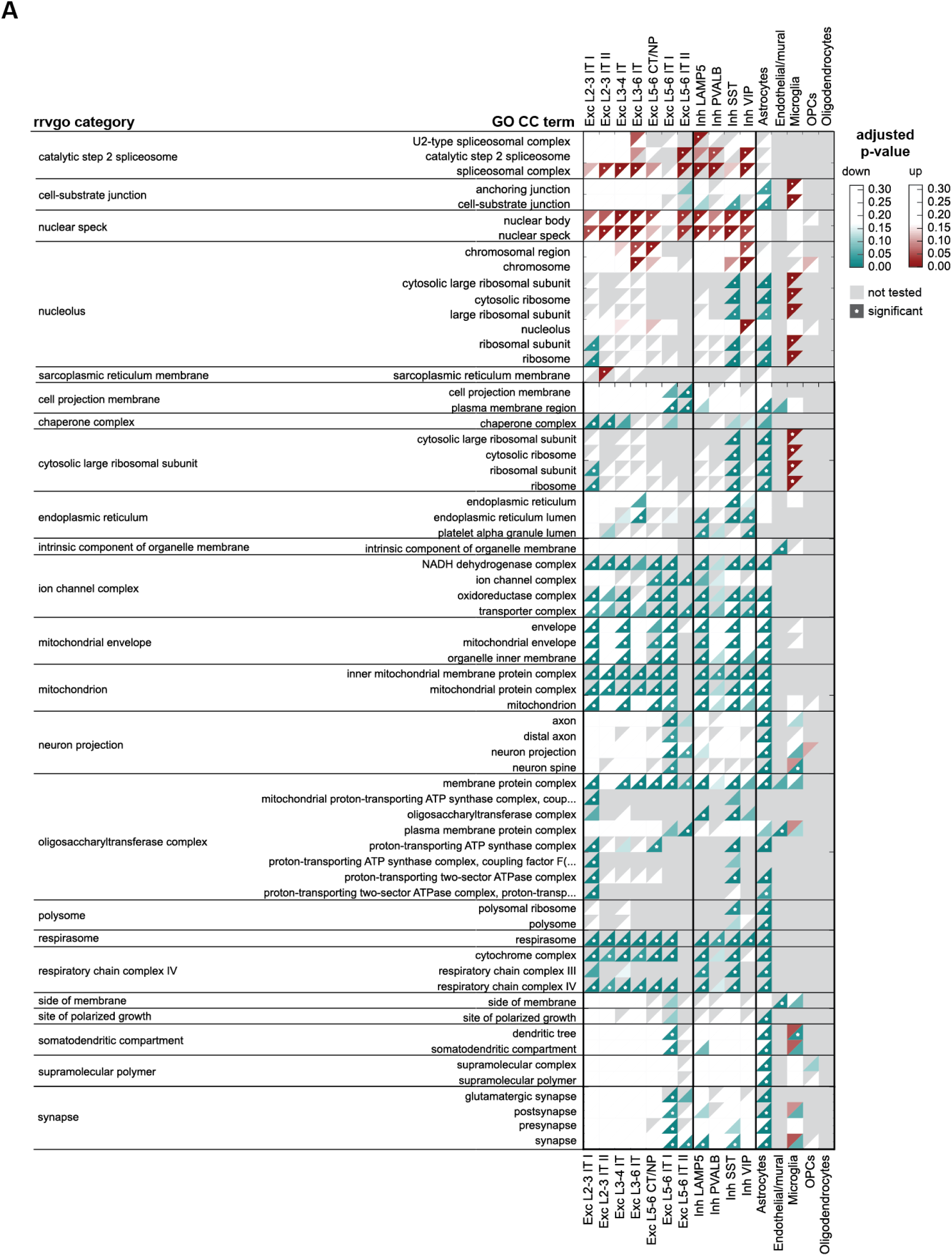
**A** GO cellular components (CC) enriched for up- (red) and down-regulated (green) DEGs. White dots indicate significant enrichment (adjusted p-value below 0.05) and gray triangles imply GSA analysis was not conducted due to insufficient overlap of DEG list with GO CC term. In total 3391 (24.9%) unique DEGs (alpha=0.3) were not mapping to any GO CC pathway and 796 (5.8%) unique DEGs (alpha=0.3) were not mapping to any GO pathway.

**Figure S11.**
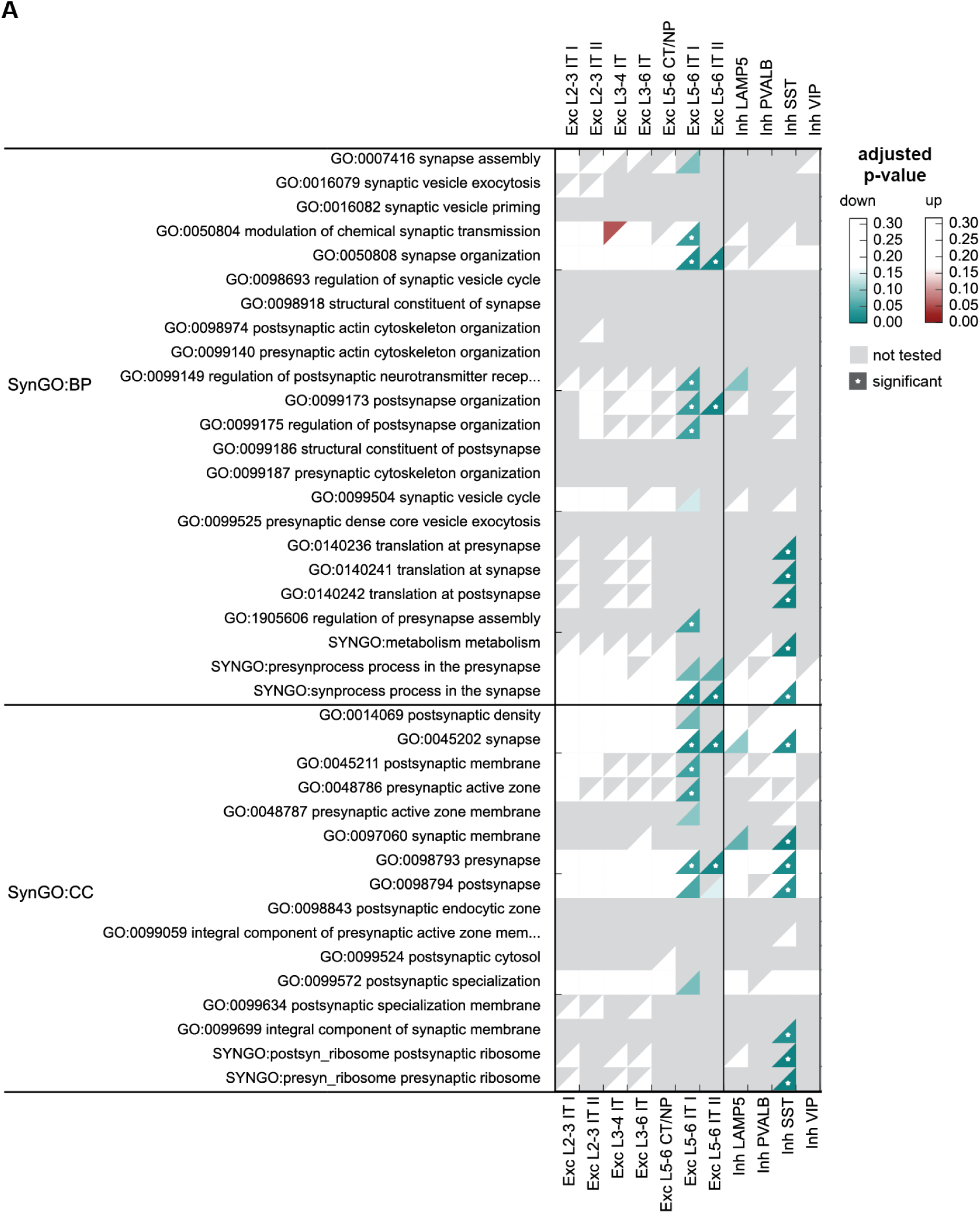
**(A)** SynGO biological processes (BP) and cellular components (CC) enriched for up- (red) and down-regulated (green) DEGs. White dots indicate significant enrichment (adjusted p-value below 0.05) and gray triangles imply GSA analysis was not conducted due to insufficient overlap of DEG list with SynGO BP term. 1147 (8.4%) unique DEGs (alpha=0.3) were overlapping with a SYNGO pathway.

**Figure S12.**
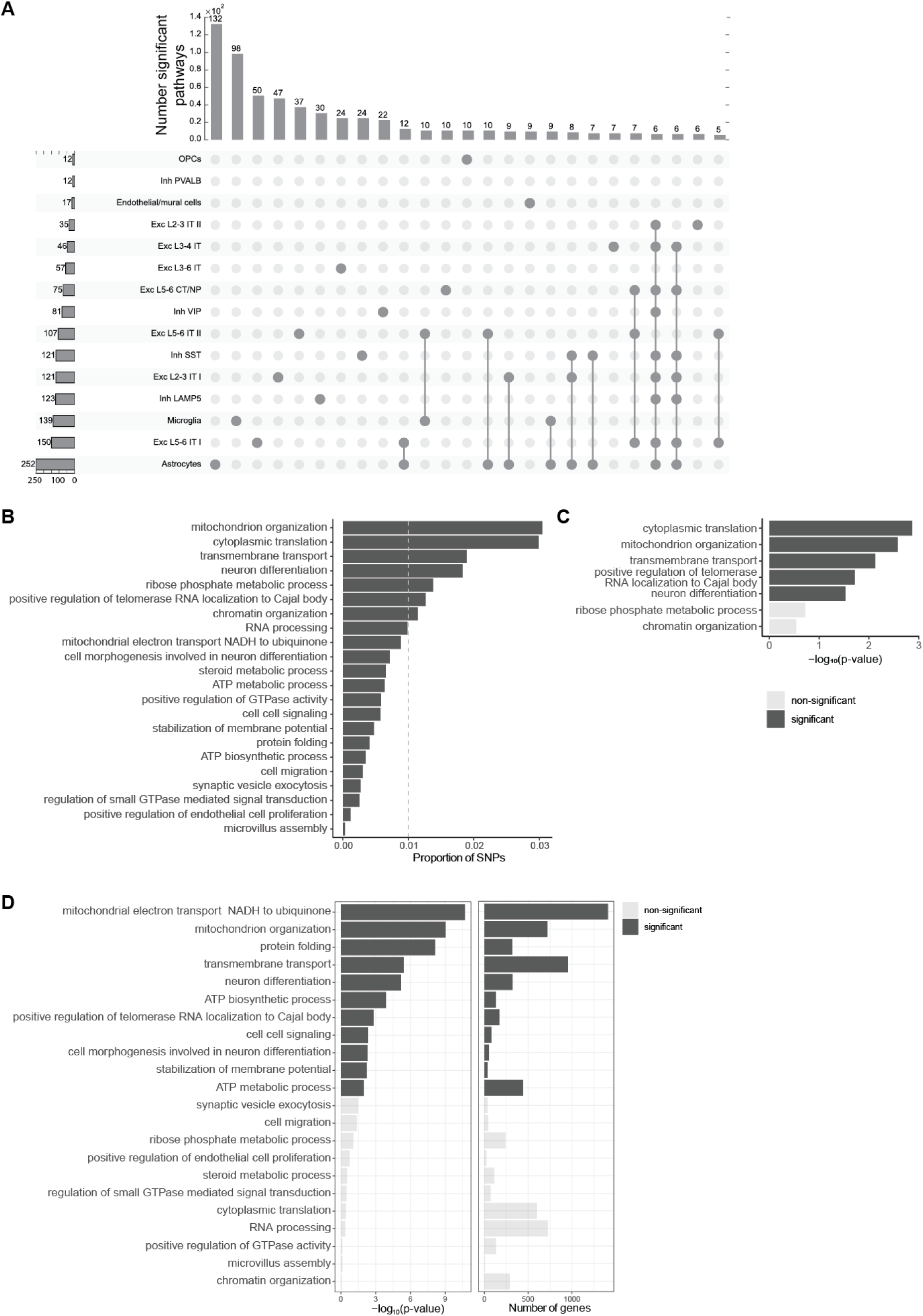
**(A)** Upset plot for significant GO terms per cell type identifying most pathways in microglia, astrocytes, excitatory IT neurons. Groups of cell types sharing less than five perturbed pathways are not shown. **(B)** Proportion of SNPs per category (in the genome-wide SNPs considered in pLDSC) is above 1% for 7 categories. **(C)** Negative log10 p-values in enrichment analysis reveals 5 significantly enriched categories. **(D)** Number of genes (right) per GO term and the significance of the enrichment (left) of mBATcombo significant genes, using negative log10 p-values as the significance measure, in agreement with LDSC implicated GO terms.

**Figure S13.**
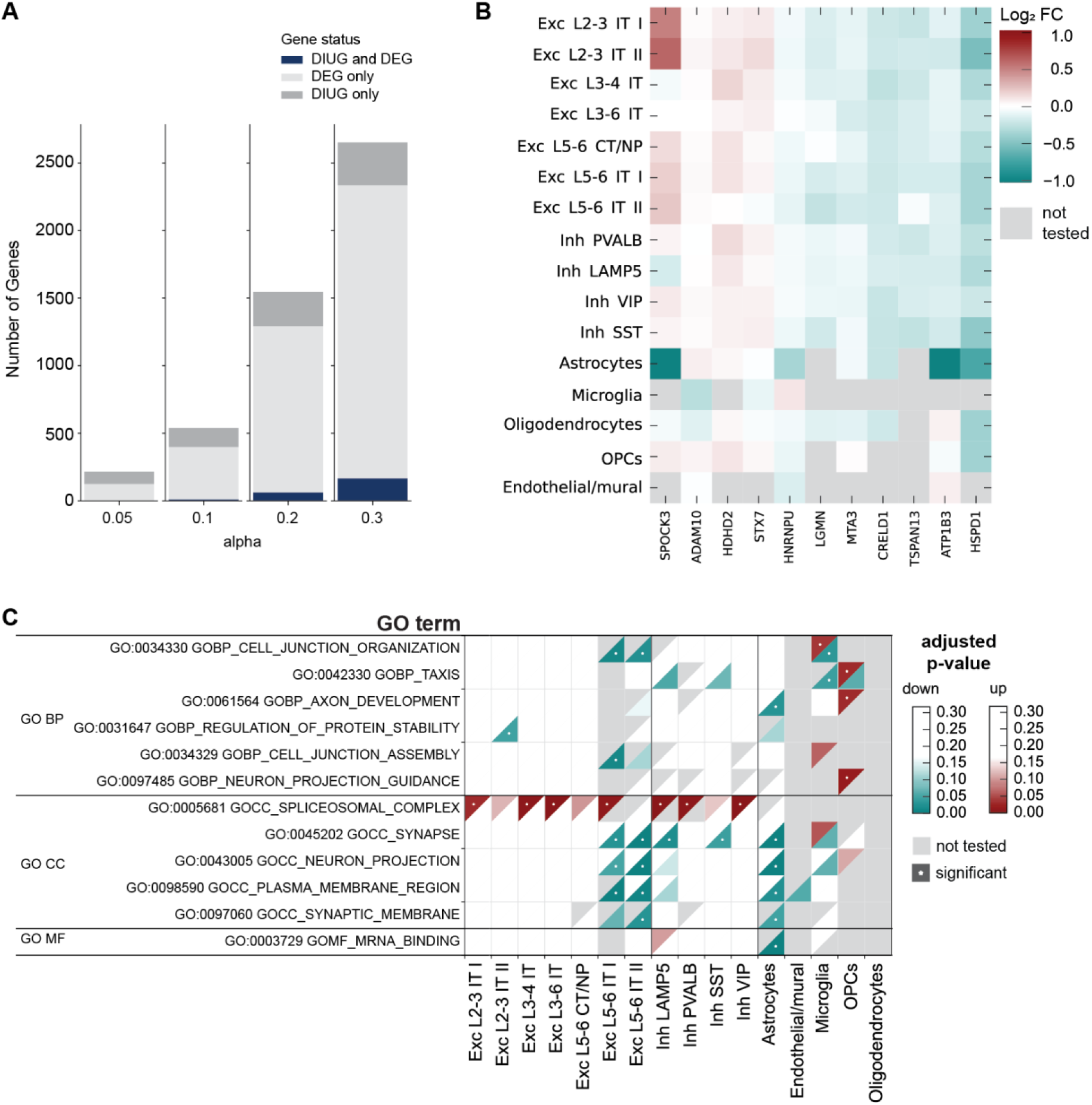
**(A)** Overlap of differentially expressed genes with differential isoform usage genes (DIUGs) for a range of alpha values indicating only little overlap of genes. **(B)** 11 DEGs overlapping for alpha=0.1 (top right) show overall rather small log_2_ fold changes in DEG analysis. **(C)** Common significant GO pathways of DIUGs and DEGs in at least one cell type are down-regulated synaptic pathways in deep layer excitatory cells and astrocytes and up-regulated spliceosomal complexes in neurons.

**Figure S14.**
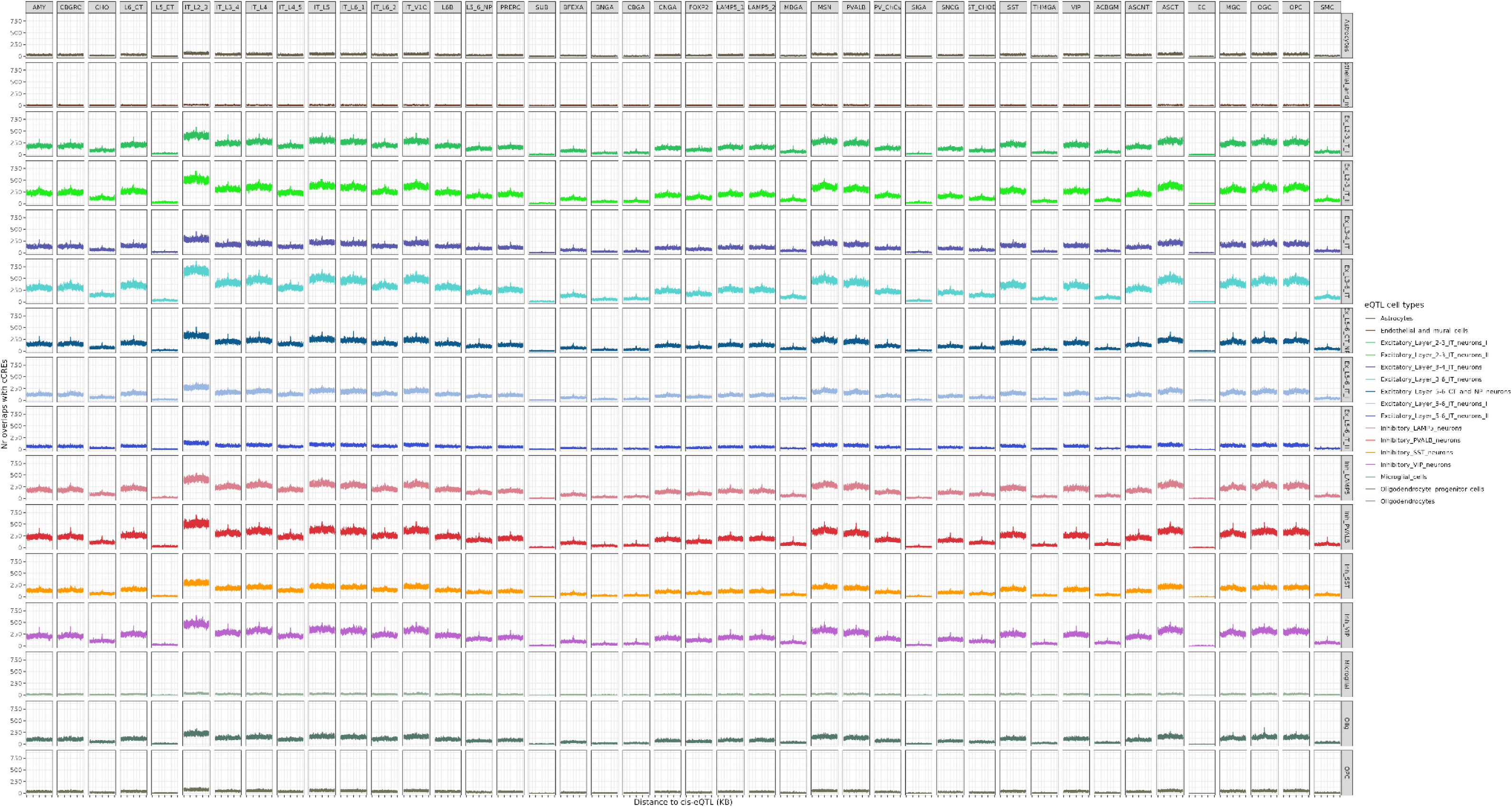
Cell type cCRE enriched around cell type eQTLs. For each eQTL, we split its 1MB flanking into 500 bp bins and counted the number of cCREs in each bin. The x-axis indicates the bins’ distance to the eQTL, and the y-axis indicates the number of cCRE observed in each bin. A peak at the eQTL position indicates an enrichment of the cCREs around eQTLs in the corresponding cell type. The names listed in the horizontal direction are the cell types of cCREs, and the names listed in the vertical direction are the cell types of the eQTLs.

**Figure S15.**
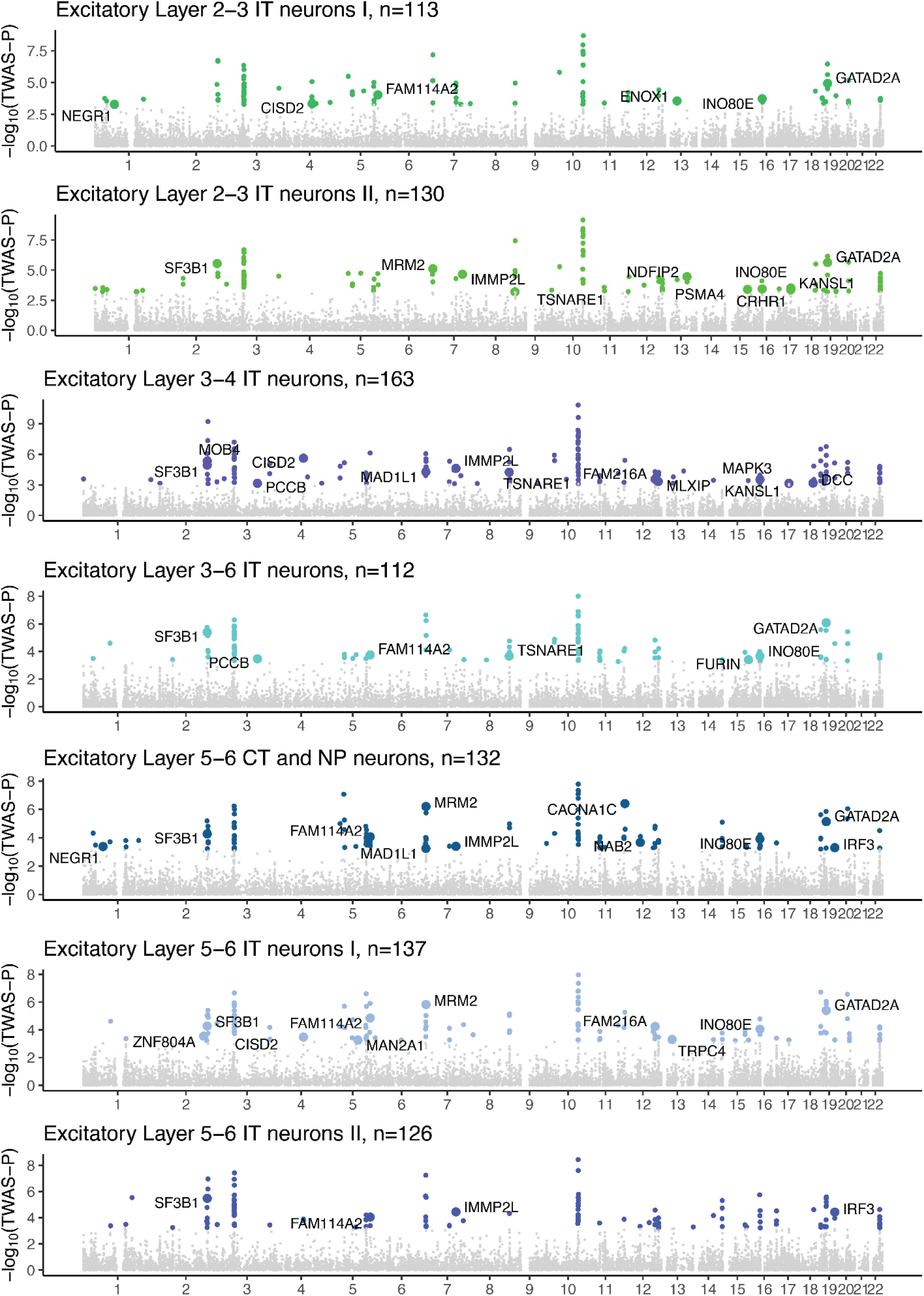
TWAS in excitatory neuronal cell types. Colored dots are significant genes (TWAS FDR≤0.05), and the enlarged, labeled dots are genes prioritized in the latest GWAS (Trubetskoy et al. 2022).

**Figure S16.**
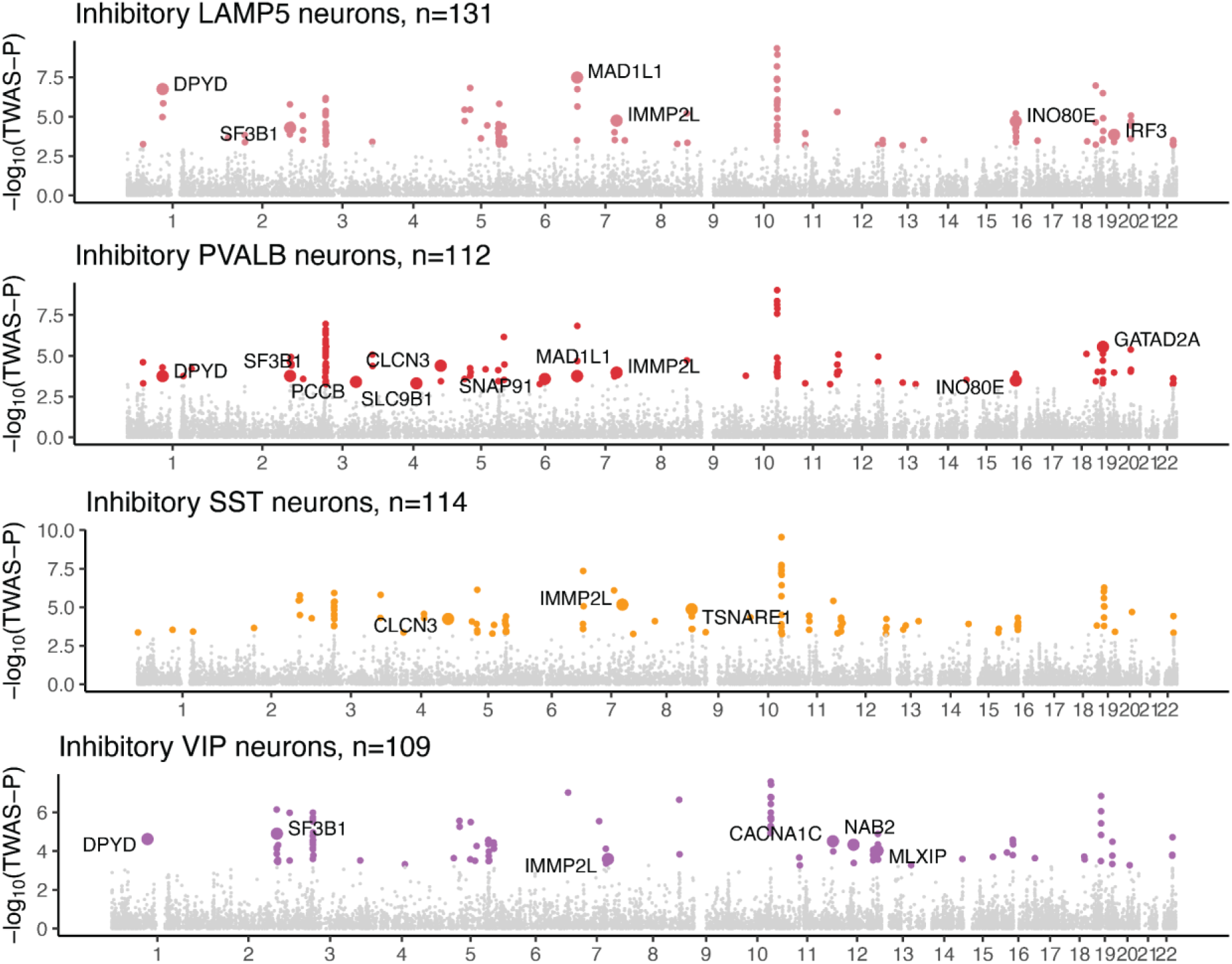
TWAS in inhibitory neuronal cell types. Colored dots are significant genes (TWAS FDR≤0.05), and the enlarged, labeled dots are genes prioritized in the latest GWAS (Trubetskoy et al. 2022).

**Figure S17.**
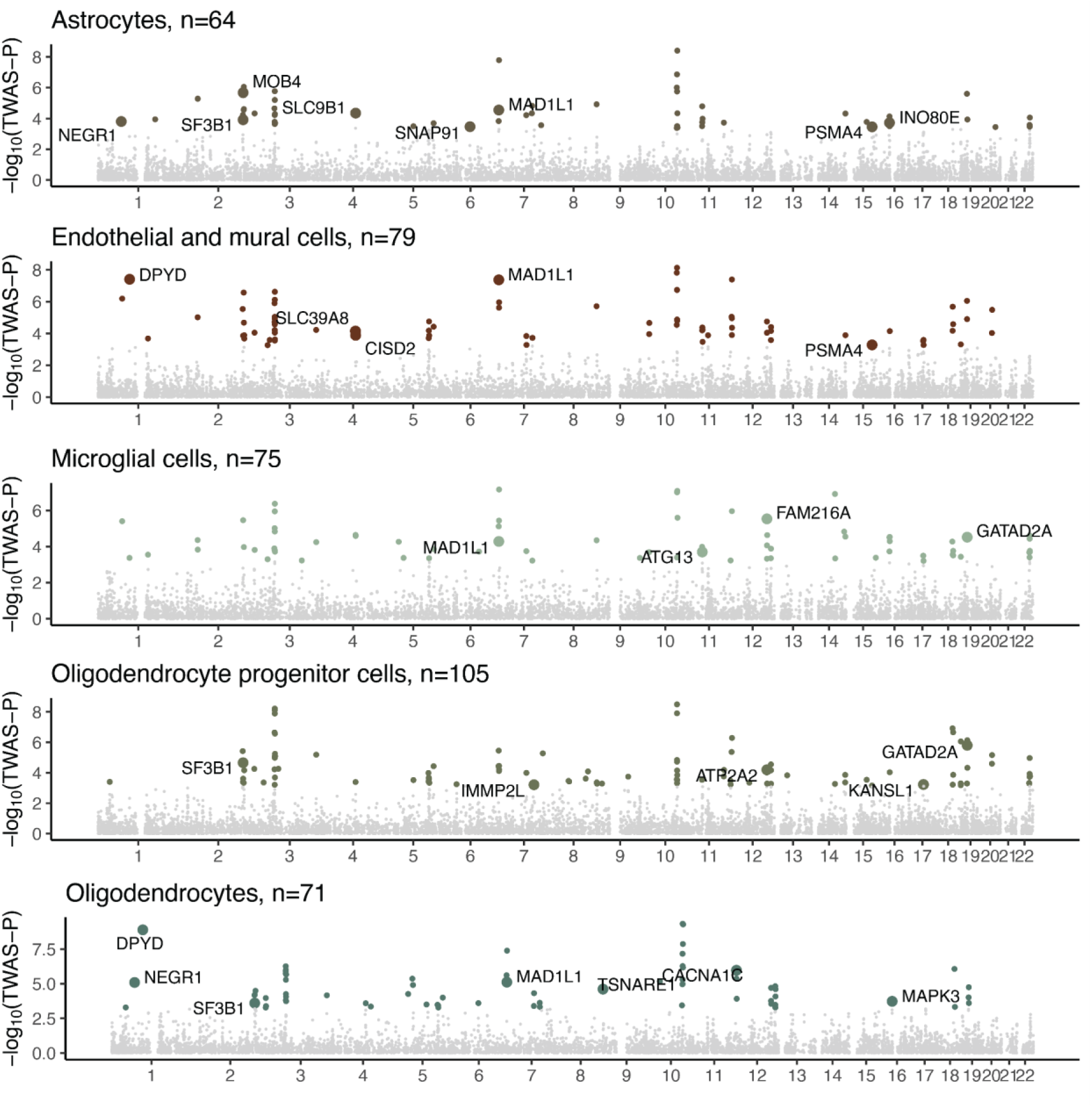
TWAS in non-neuronal cell types. Colored dots are significant genes (TWAS FDR≤0.05), and the enlarged, labeled dots are genes prioritized in the latest GWAS (Trubetskoy et al. 2022).

**Figure S18.**
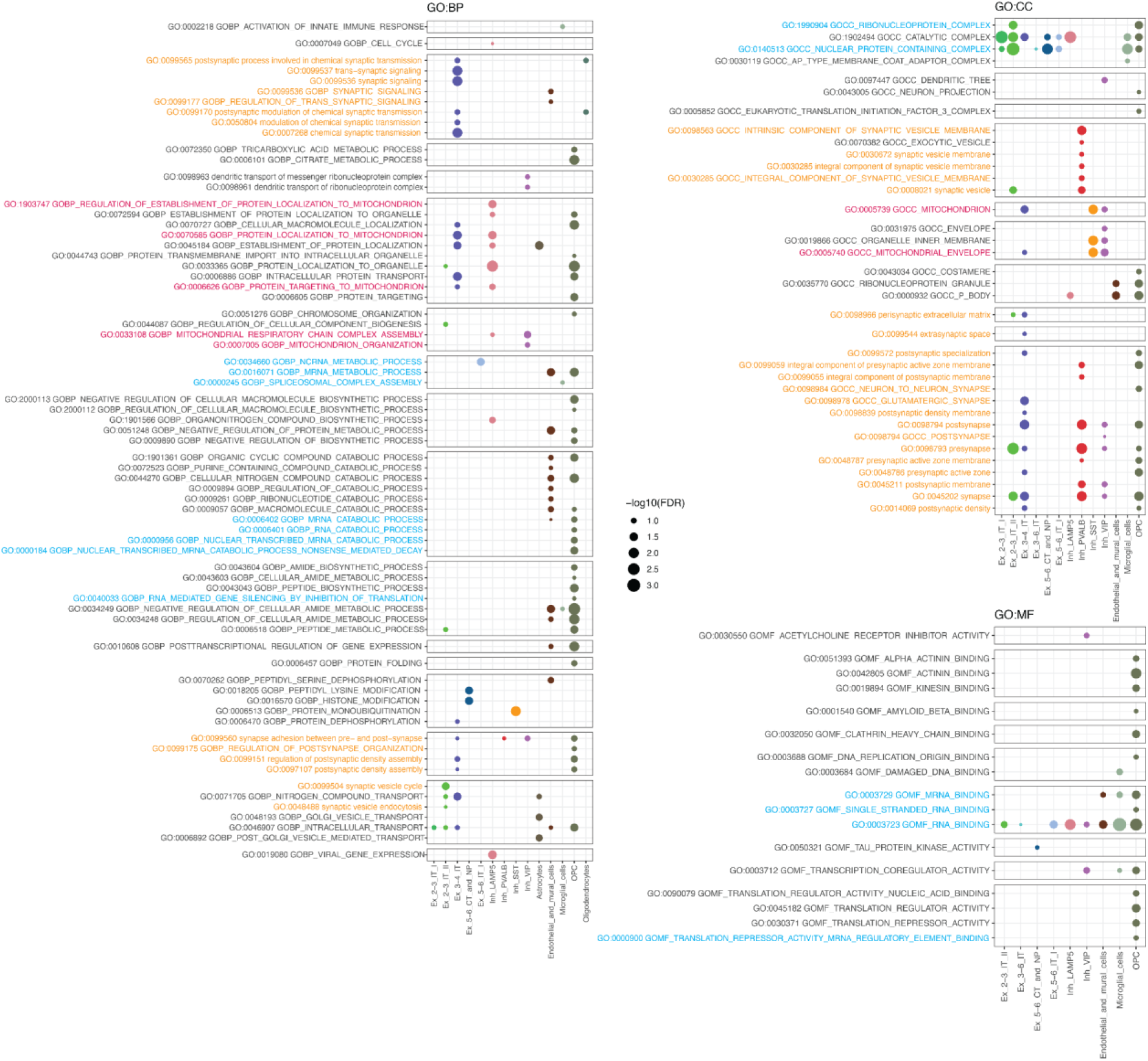
GO pathways enriched in TWAS genes per cell type, according to hypergeometric tests (TWAS-FDR≤0.3), including multiple testing corrections in each database source, i.e., GO:BP, GO:CC, and GO:MF. Plotted are pathways with enrichment FDR≤0.15, text highlighted in blue are RNA-processing pathways, in red mitochondrial/ATP pathways, and in orange synaptic pathways.

**Figure S19.**
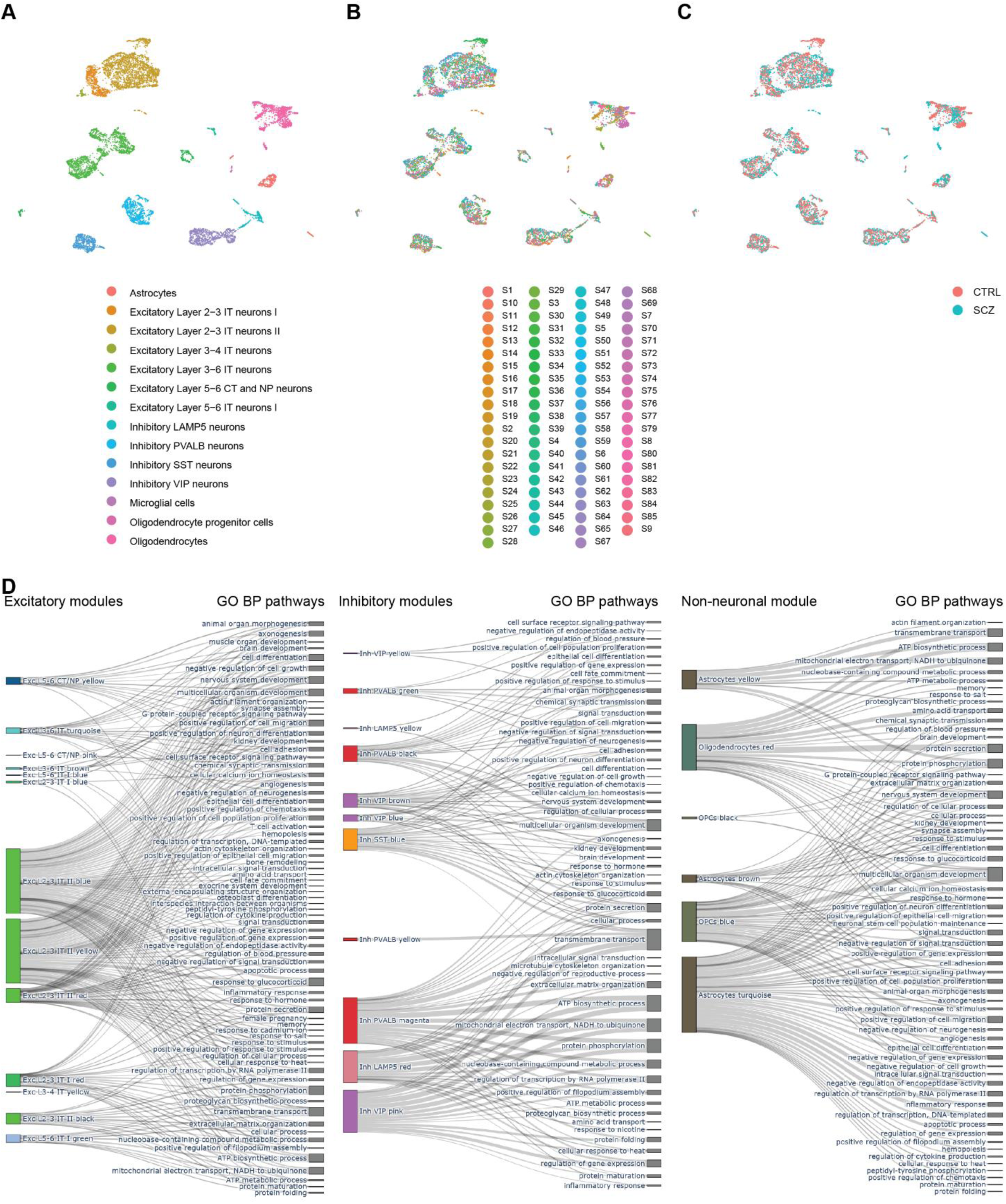
Metacells constructed for gene regulatory network inference highlighting **(A)** cell types, **(B)** donors and **(C)** group. **(D)** Cell type specific modules significantly implicated with schizophrenia and sorted by their cell type class excitatory (left), inhibitory (middle), non-neuronal (right) mapped to their significant (alpha=0.05, FDR corrected by number of pathways in respective group) GO BP parent term.

**Figure S20.**
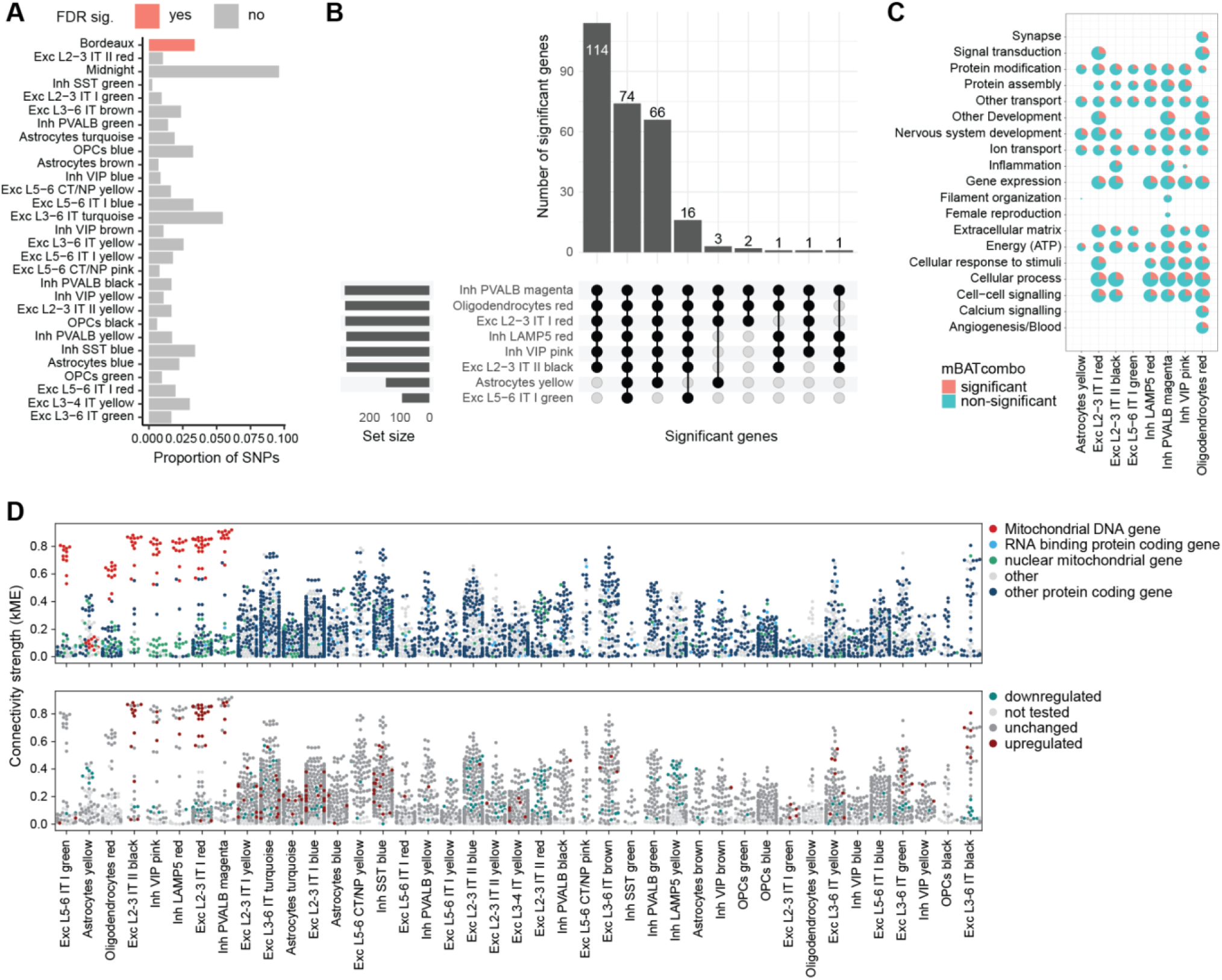
**(A)** Proportion of SNPs per module (group) is highest for midnight modules. **(B)** Overlap of significantly enriched genes per bordeaux module. **(C)** Percentage of genes significantly enriched for SCZ risk per function and bordeaux module. **(D)** Connectivity strength of each gene per module significantly changed in schizophrenia highlighting each gene’s type (upper row) and (de-)regulation status in DEG analysis (lower row).

**Figure S21.**
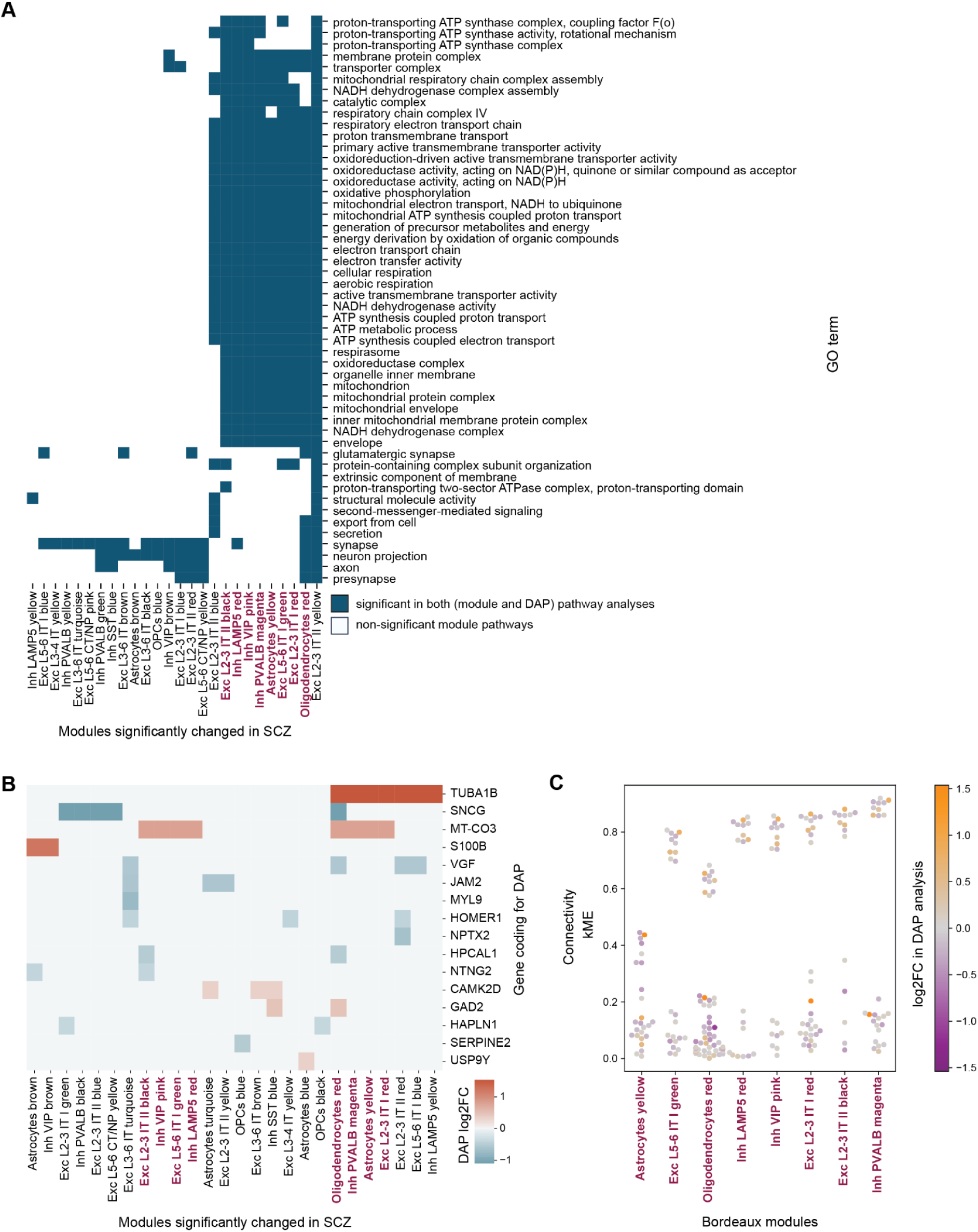
**(A)** Significant GO pathways of schizophrenia-relevant modules overlapping with GO pathways of proteomics are predominantly mitochondrial, energy and synaptic pathways in bordeaux modules. **(B)** Genes in modules that are coding for proteins and their log_2_ fold changes in differentially abundant protein (DAP) analysis. **(C)** Connectivity of bordeaux modules per gene coding for proteins and their log_2_ fold changes in DAP analysis. Genes with high connectivity are coding for rather more abundant than less abundant proteins.

**Figure S22.**
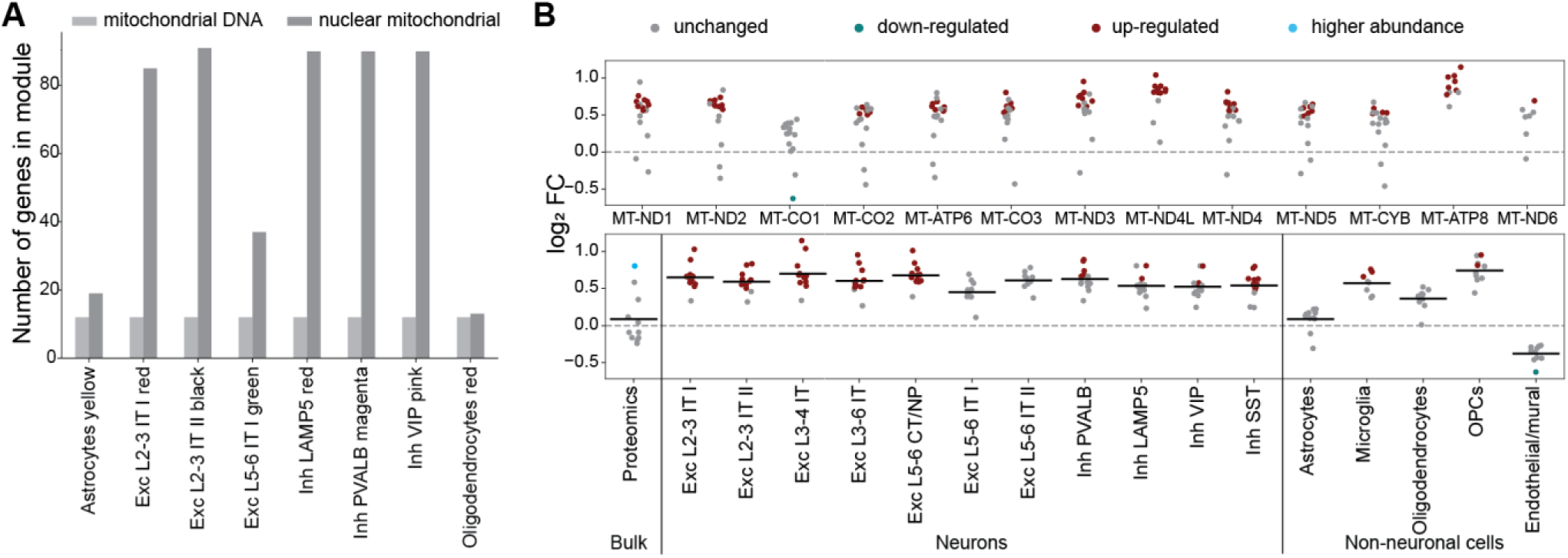
**(A)** Number of mitochondrial DNA and nuclear mitochondrial genes per bordeaux module is particularly high in neuronal modules. All bordeaux modules contain mitochondrial DNA genes. **(B)** Mitochondrial DNA genes have increased expression in the schizophrenia group in almost all cell types and are mostly unchanged in bulk proteomics. Endothelial and mural cells show a slightly decreased expression in mitochondrial DNA genes.

**Table T1:** Basic donor information T1_basic_donor_information.xlsx

**Table T2:** Quality metrics per sample T2_quality_metrics_per_sample.xlsx

**Table T3:**
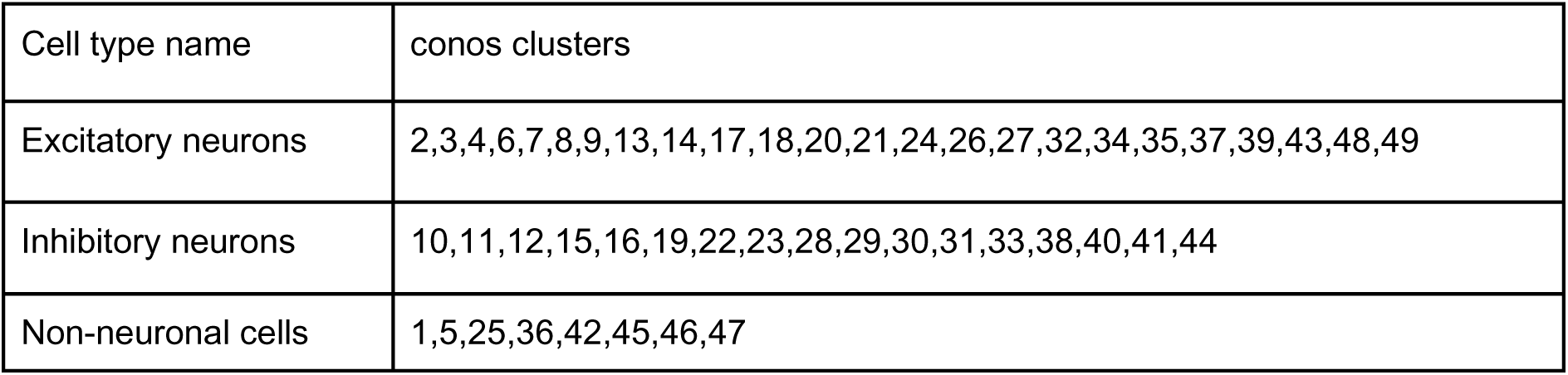
Cell classes and their corresponding conos clusters.

**Table T4:**
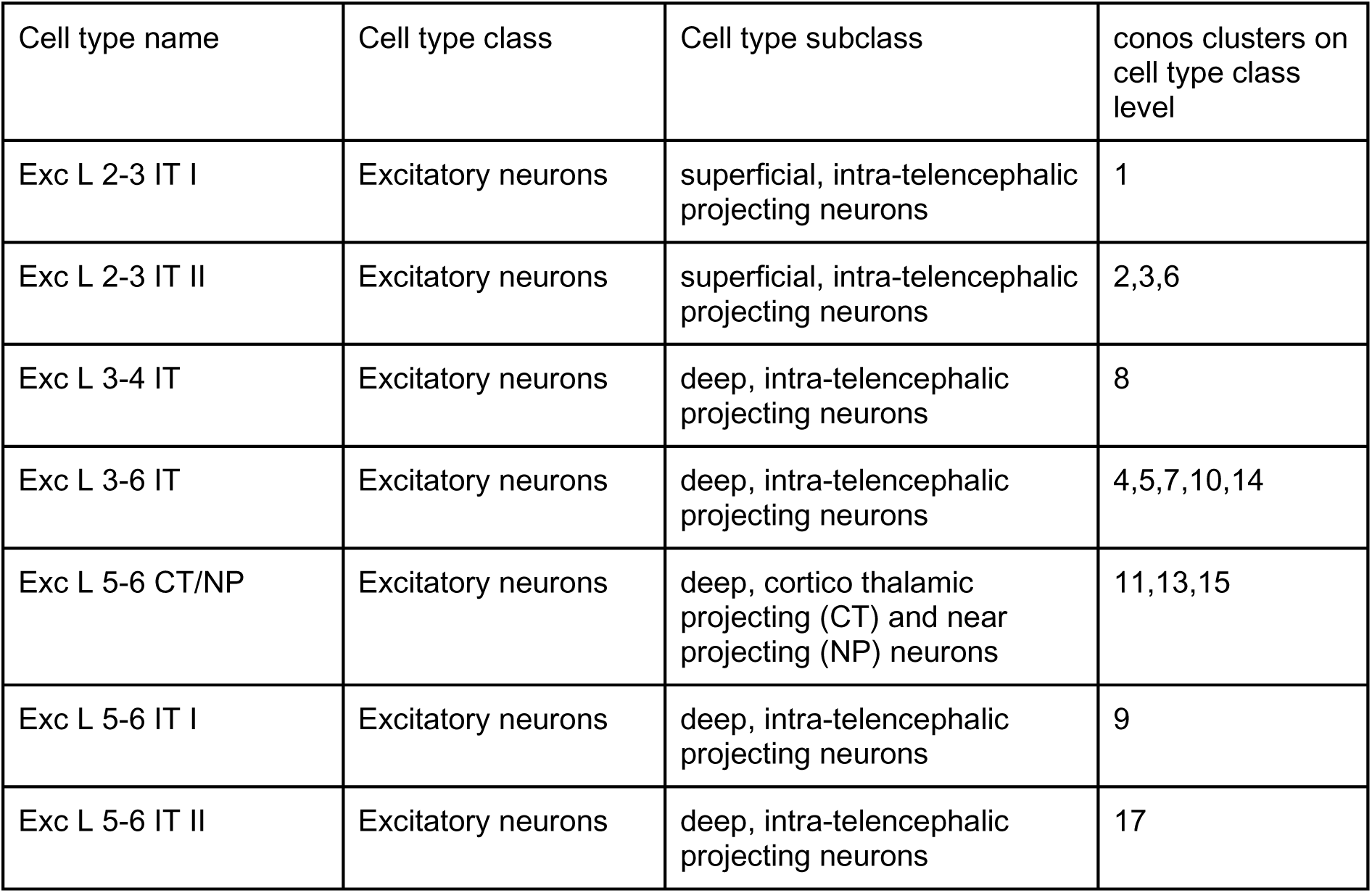
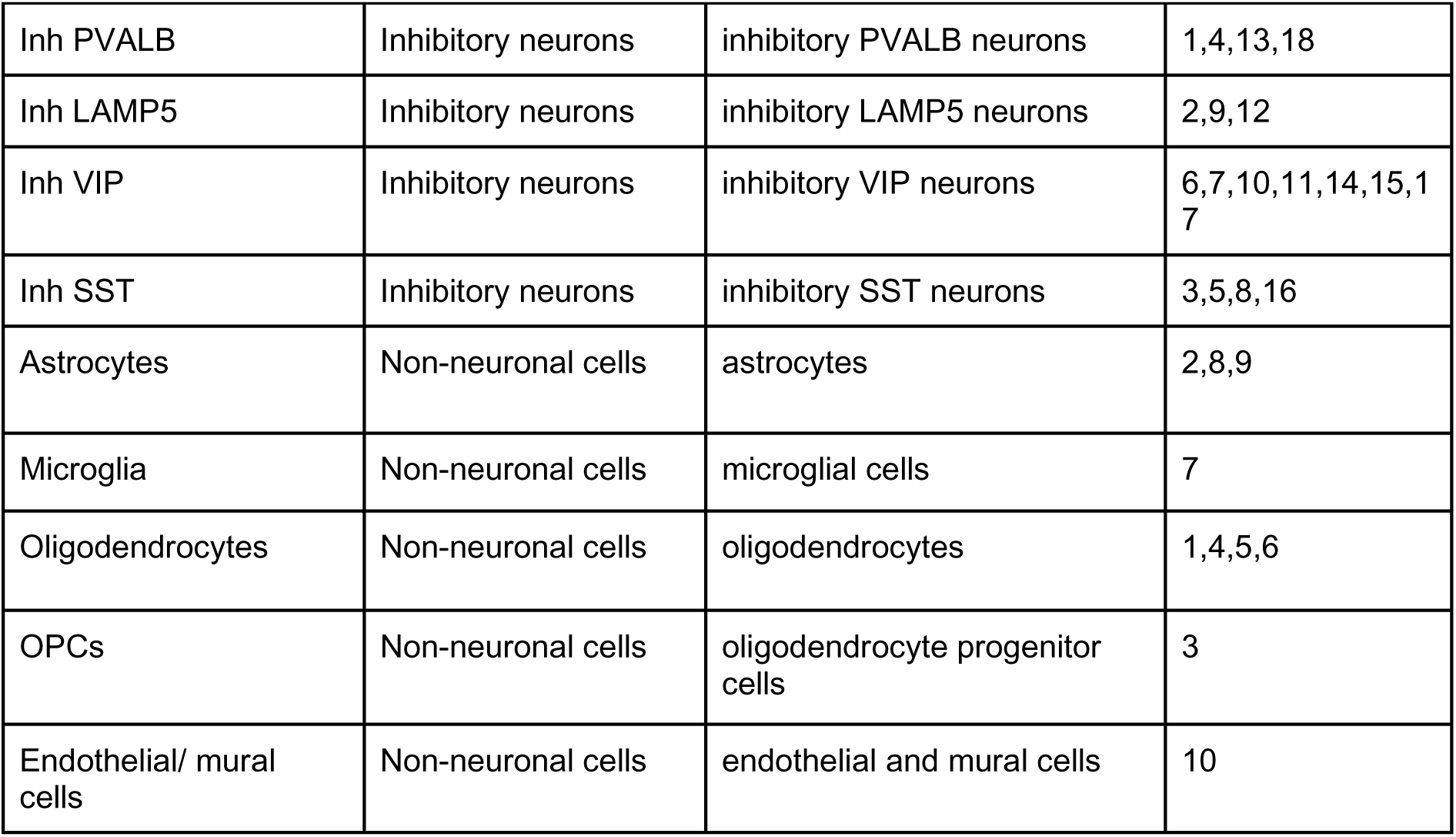
Cell type names and their corresponding conos clusters.

**Table T5:** Up- and down-regulated DEGs per cell type.

T5_DEGs_per_cell_type.csv

**Table T6:** Pathways including GO terms with their within subgroup adjusted p-values for up- and down-regulated DEGs per celltype.

T6_sign_pathways_per_cell_type.xlsx

**Table T7:** Sparsity result for a range of parameter values for metacell calculation.

T7_metacell_sparsity_result.csv.

**Table T8:** GO BP terms per module.

T8_pathway_results_modules.xlsx

**Table T9:** GO BP term category mapping.

T9_Term_category_mapping.xlsx

**Table T10:**
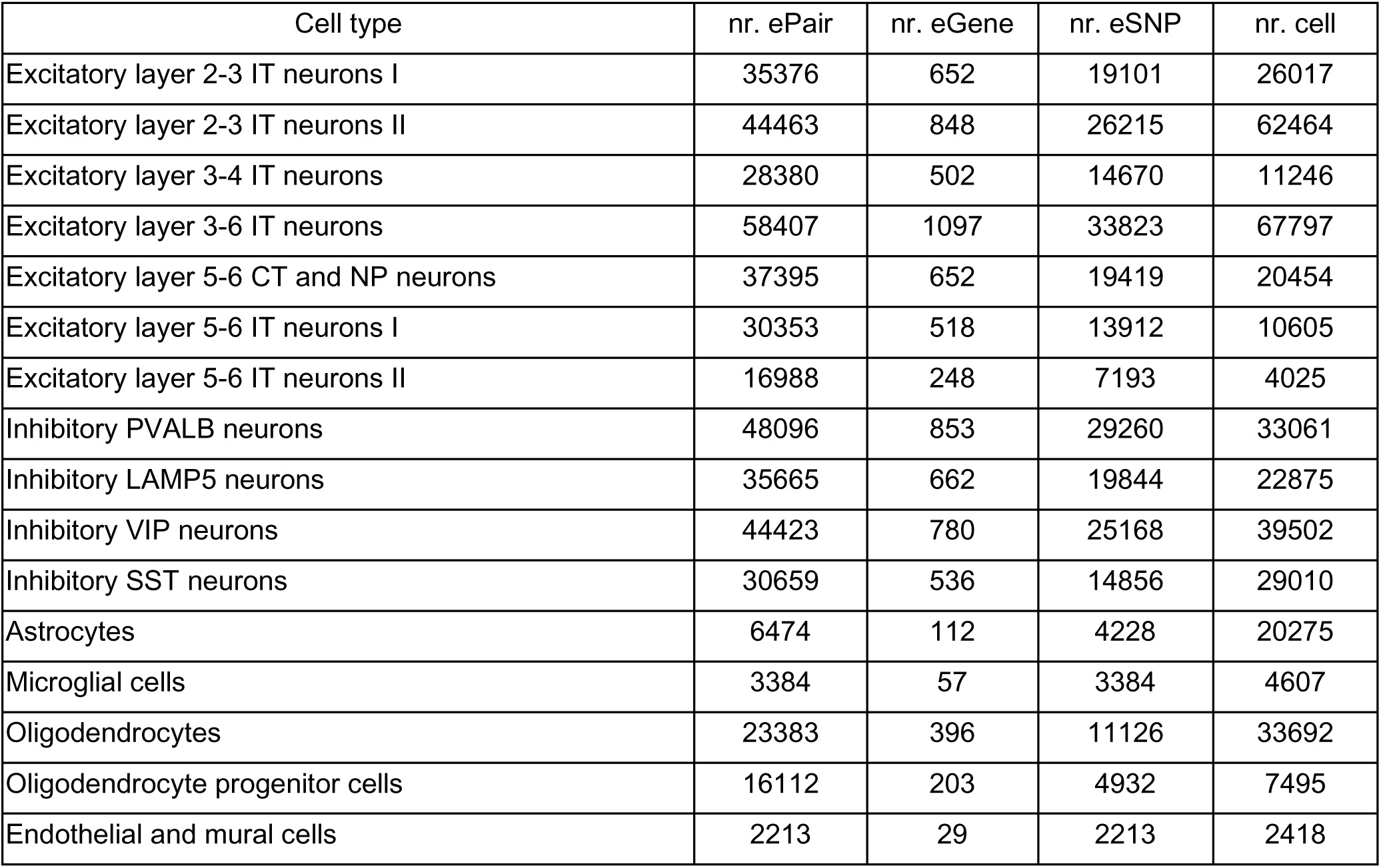
Numbers of eQTL pairs per cell type.

**Table T11:**
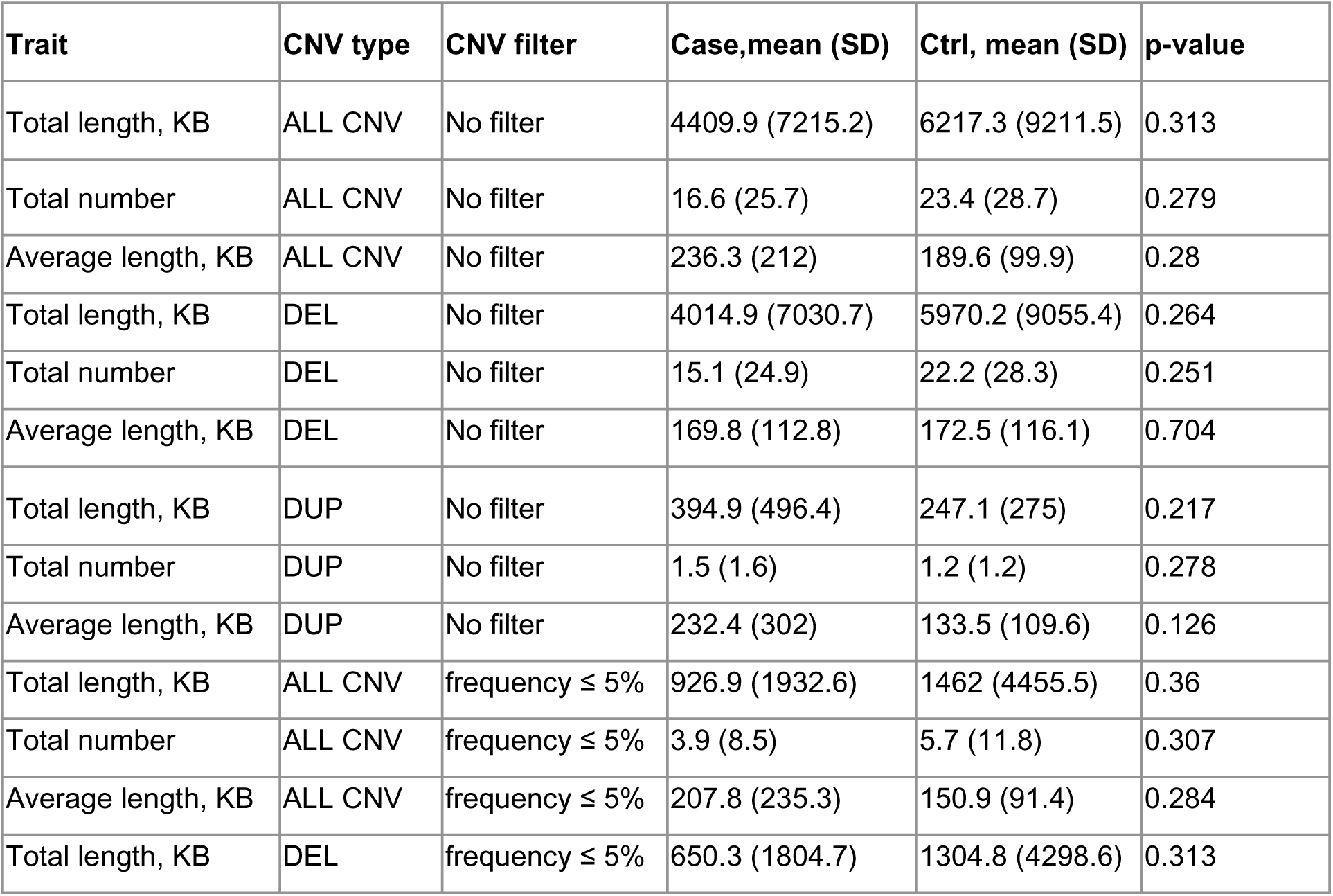
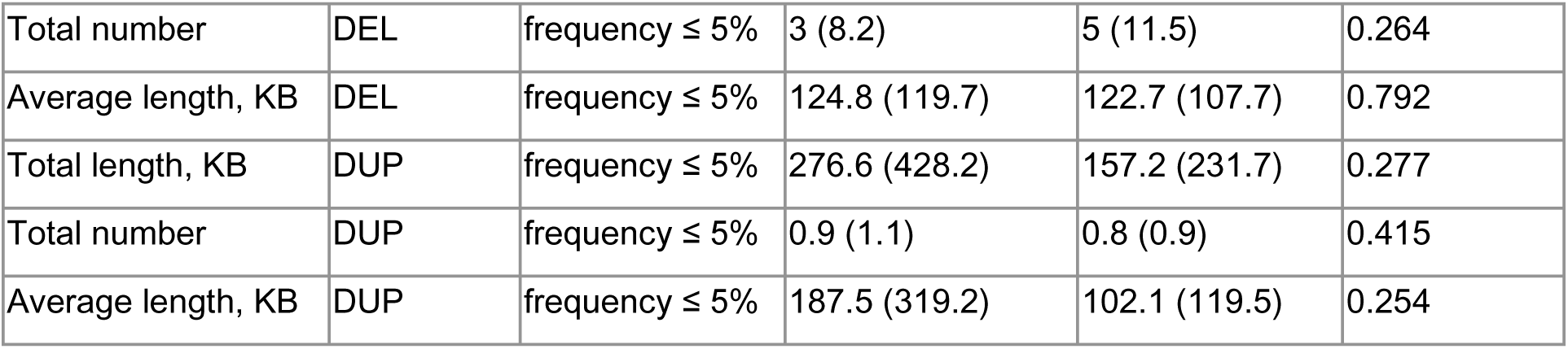
CNV burden test.

**Table T12:**
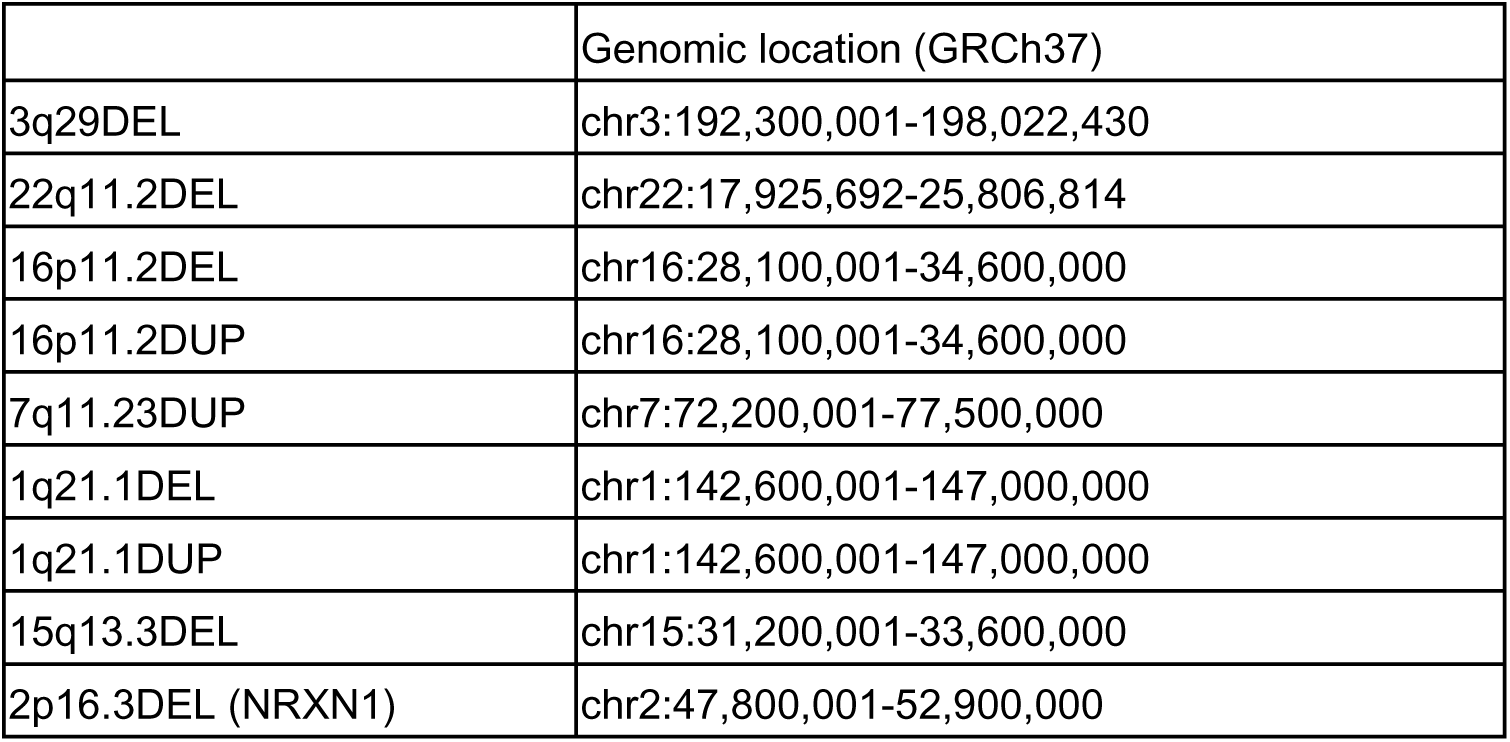
CNVs related to schizophrenia.

**Table T13:** Full TWAS results at cell type level.

T13_TWAS_full_results.tsv

**Table T14:** mBATcombo results for bordeaux module genes.

T14_mBATcombo_bordeaux_module_genes.tsv

## Data availability

The source data and processed datasets that support the findings of this study are in the process of being made available through the European Genome-Phenome Archive (EGA, https://ega-archive.org). Table files can be accessed through DOI:10.5281/zenodo.15025718. The GeneMatrix resource is available online at figshare https://figshare.com/articles/dataset/geneMatrix/13335548.

## Code availability

All original code is available at https://github.com/Hjerling-Leffler-Lab/integrative_omics_schizophrenia.

## Acknowledgements

We gratefully acknowledge the support of multiple funders to this work: Swedish Research Council (Vetenskapsrådet, award D0886501, PFS), European Research Council (SCHIZTYPE, award 819540, JHL) Swedish Research Council (Vetenskapsrådet, award 2018-00799, JHL), Hjärnfonden (Swedish Brain Foundation, award FO2021-0210, JHL), StratNeuro (SY), Brain & Behavior Research Foundation (NARSAD Young Investigator Grant 2022-31182, JS), Swedish Research Council (Vetenskapsrådet, award 2022-00242, RZ), IDeA COBRE award from the National Institute of General Medical Sciences (P30 GM103328, CAS).

The authors gratefully acknowledge contributions of the Cuyahoga County Medical Examiner’s Office and the families of the deceased. The efforts of Drs. James Overholser, George Jurjus and Herbert Y. Meltzer, and Lesa Dieter and Gouri Mahajan in the psychiatric assessments and tissue processing are also acknowledged. FACS isolation was performed with support from Biomedicum Flow Cytometry Core Facility (BFC). We acknowledge Anke A. Dijkstra and Remco V. Klaassen for technical assistance and performing mass spectrometry experiments. The authors acknowledge support from the National Genomics Infrastructure in Stockholm funded by Science for Life Laboratory, the Knut and Alice Wallenberg Foundation and the Swedish Research Council, and NAISS/Uppsala Multidisciplinary Center for Advanced Computational Science for assistance with massively parallel sequencing and access to the UPPMAX computational infrastructure. Computational resources, data handling and support were enabled by the National Academic Infrastructure for Supercomputing in Sweden (NAISS), hosted by UPPMAX and PDC, partially funded by the Swedish Research Council through grant agreement no. 2022-06725, and by the University of North Carolina at Chapel Hill and the Research Computing group.

## Author Contributions

Conceptualization (JHL, PFS), Data Curation (LB, SY, RK, JML), Formal Analysis (LB, SY, ML, JML, HF, RK, JW, JS), Funding Acquisition (JHL, PFS, ABS, PGR), Investigation (FM, LB, JML, FK, MV, CS, AD), Methodology (LB, SY, RK, RZ, JW, JS, ML), Project Administration (FM, AMÖ, JHL, PFS), Resources (JHL, PFS, ABS, PGR, CS, AD), Software (LB, SY, HF, JML), Supervision (JHL, PFS, ABS, PGR), Validation (LB, SY, HF, RK, JW, JS), Visualization (LB, SY, JML, JHL, PFS), Writing - Original Draft Preparation (LB, SY, PFS, JHL), Writing - Review and Editing (all authors).

